# A proteomic signature for prognostic stratification in amyotrophic lateral sclerosis

**DOI:** 10.64898/2026.07.20.26358511

**Authors:** David G Lester, Paolo Piazza, Elizabeth Dellar, Prerak Desai, Hadar Klimovski, Christos V Chalitsios, Marcel Weinreich, Elham Alhathli, Andrew Strange, Dganit Melamed-Kadosh, Tamar Ziv, Pamela J Shaw, Arie Admon, Vivian Drory, Andrea Malaspina, Johnathan Cooper-Knock, Iddo Magen, Eran Hornstein, Armina Omole, Guhan Nagappan, Avigail Taylor, John A Todd, Kevin Talbot, Martin R Turner, Alexander G Thompson

## Abstract

Biochemical predictors of survival are essential to control heterogeneity and identify associations of disease progression in amyotrophic lateral sclerosis (ALS). We performed high-depth Proximity Extension Assay proteomics using longitudinal serum (*n*=851) and CSF (*n*=244) samples from 426 people with ALS, replicated in a further 460 patients. Cox regression identified five CSF proteins, including tropomyosin 3 (TPM3), and 57 serum proteins, including neurofilament light chain (NEFL) and peripherin, associated with survival. Lasso regression identified an eight-protein serum panel, including NEFL, peripherin, EDA2R and CALCA — reflecting both disease extent and progression rate — that improved survival prediction beyond clinical parameters and NEFL: external validation *c*-statistic 0.78 versus 0.74 (Δ 95% CI 0.00-0.08), *p*=0.066; 1-year AUC 0.89 versus 0.84 (0.00-0.11), *p*=0.047. Joint modelling identified survival-associated trajectories of serum EDA2R and CALCA, highlighting their potential to measure disease progression. These findings identify diverse serum proteins that enhance survival stratification and are associated with disease progression in ALS.

## INTRODUCTION

Amyotrophic lateral sclerosis (ALS) is a degenerative disorder of the corticomotoneuronal network, characterised by progressive motor neuron loss leading to muscle weakness and respiratory failure (Brown et al., 2017). ALS is typically life-shortening, with a median survival of 30 months from symptom onset (Brown et al., 2017). However, disease progression is highly variable, with up to 10% of people with ALS surviving beyond 10 years (Chiò et al., 2011). This heterogeneity reflects a complex and poorly understood pathobiology of disease progression, involving molecular, systemic and neural connectome factors (Brown et al., 2017). Riluzole, the only widely approved treatment for sporadic ALS, extends life expectancy by an average of only two to three months, highlighting the need to better understand the factors determining disease progression (Miller et al., 2012).

Variation in ALS prognosis presents a major challenge for the development of effective disease-modifying therapies (Mitsumoto et al., 2014). Prognostic models grounded in the biology of disease progression are therefore urgently needed (Van Den Berg et al., 2019). However, limited understanding of the mechanisms underlying survival heterogeneity means that established prognostic models largely rely on clinical observations remote from disease biology, and which are susceptible to non-linearity, ceiling and floor effects, and inter-rater variability (Westeneng et al., 2018; Van Eijk et al., 2021).

Incorporating biochemical data into prognostic models may offer a route towards precision medicine in ALS. To date, this approach has focused predominantly on neurofilament light chain (NEFL) protein levels measured in cerebrospinal fluid (CSF) and blood, which reflect the rate of axonal degeneration (Gaiani et al., 2017; Benatar et al., 2020; Thompson et al., 2022; Benatar et al., 2024). Deeper integration of more complex biochemistry may capture additional biological processes associated with disease progression, providing objective biomarkers for prognostic modelling and therapeutic trials.

We combined high-depth biofluid proteomic and clinical data from multiple large, well-characterised longitudinal cohorts to identify prognostic biomarkers in ALS and evaluate their performance relative to clinical predictors and NEFL levels. We further applied functional enrichment and genetic causal analyses to investigate the identified prognostic candidates with disease progression.

## METHODS

### Participants and sampling

People with ALS were diagnosed by neurologists specialised in motor disorders, based on upper and lower motor neuron (UMN; LMN) dysfunction in at least one region or LMN dysfunction in at least two regions, with investigations excluding other causes. For the discovery cohort, participants attending the Oxford MND Care Centre, John Radcliffe Hospital, Oxford, were recruited into six research ethics committee (REC)-approved sample collection cohorts between 2009 and 2023 (**Supp. Table 1**). All participants provided informed consent. There were no additional exclusion criteria, although lumbar puncture was not offered to individuals taking clopidogrel or anticoagulants.

Peripheral blood and CSF were collected in serum separator and polypropylene tubes, respectively, and processed according to consensus biomarker guidelines (Teunissen et al., 2009). Samples were centrifuged at 2300 x *g* for 10 minutes at 4°C, aliquoted into polypropylene tubes and frozen within 2 hours of collection. AMBRoSIA cohort samples were stored in vapour phase nitrogen (VPN; -180°C), whereas samples from the remaining cohorts were stored at -80°C. AMBRoSIA and PRoSNeB samples had not previously been thawed, whereas samples from the remaining cohorts had been thawed once before (**Supp. Tables 2-3**).

### Clinical measurements

Clinical measurements at sampling included symptom onset date and site (bulbar or non-bulbar), ALS Functional Rating Scale-Revised (ALSFRS-R), Edinburgh Cognitive and Behavioural ALS Screen (ECAS), neurologic examination, body mass index (BMI) and forced vital capacity (FVC, % predicted). Genetic testing was performed through standard clinical pathways. Progression rate was calculated as (48-ALSFRS-R)/ symptom duration (months). Clinical UMN and LMN burden was assessed across five body regions using hyperreflexia, wasting and fasciculation (score range -10 to +10, representing most flaccid to most spastic; Gao et al., 2022). Date of death or censoring was ascertained from electronic health records in 2024.

### Oxford discovery Proximity Extension Assay

Samples were thawed on ice and pipetted onto 96-well plates using layouts designed to distribute biological and technical factors within and across plates (Taylor et al., 2025). CSF and serum proteomic profiling were performed using Olink Explore 3072 (v3.8.1; Thermo Fisher Scientific) and Explore HT (v6.7.2), respectively, at the Centre for Human Genetics (CHG), University of Oxford. Proximity Extension Assay (PEA) uses oligonucleotide-tagged antibody pairs that, upon binding their target protein in solution, bring the oligonucleotides into proximity for enzymatic extension, generating DNA barcodes. These are amplified and quantified using next-generation sequencing (Illumina NovaSeq) to generate relative, log_2_-transformed Normalised Protein eXpression (NPX) values (Wik et al., 2021).

Three internal controls were spiked into each sample to monitor background signal, DNA polymerase activity and sequencing performance. External sample and negative controls were used to assess intra- and inter-plate assay coefficient of variation (CV), and limit of detection (LOD), respectively. LOD was defined as 3 SD above the median negative control value for each plate. Dilution series were used mitigate the hook effect and extend the reliable detection range. Followiing plate layout optimisation, intensity normalisation was applied to equalise median assay values across plates.

### Quality control

Median (Q1-Q3) intra- and inter-plate CVs were 7.1% (3.6%-15.5%) and 11.5% (7.5%-19.8%) for CSF, and 23.7% (12.5%-41.4%) and 21.2% (12.4%-34.7%) for serum, respectively. The median proportion of samples below the assay-specific LOD per protein was 16.9% (0%-71.6%) for CSF and 87.8% (4.9%-100%) for serum (**Supp. Fig. 1**). Samples were excluded according to predefined criteria (>5 SD from the mean for PC1, PC2, mean sample NPX or NPX IQR), resulting in the removal of no CSF samples and six serum samples before further analysis (**Supp. Fig. 2**; Sun et al., 2023). Six CSF and eight serum assays had quality control (QC) warnings but were retained following inspection of raw count histograms (**Supp. Fig. 3**). In accordance with standard PEA analysis pipelines, values below LOD were retained for analysis. Associations between biological and technical factors and PC1-10 were assessed using linear models and *R*^2^. PEA measurements were compared to existing Meso Scale Discovery (MSD) electrochemiluminescence and ELISA measurements from the same samples, as previously described (Thompson et al., 2022).

### Tel Aviv mass spectrometry replication

Patients with ALS were assessed at Tel Aviv Sourasky Medical Center, Israel, between 2006 and 2019 (Klimovski et al., 2026). First-visit serum samples (*n*=349) were analysed using data-independent acquisition mass spectrometry (MS). Samples were collected within an IRB-approved protocol (0245-10-TLV) by informed consent and stored at -80°C. Protein abundances underwent QC filtering, log_2_ transformation, normalisation and batch correction using linear modelling. These data were used for significant serum protein Cox replication, with survival defined by time to death or tracheostomy.

### Sheffield PEA replication

External serum PEA replication samples were obtained from patients with ALS attending the Sheffield MND Care Centre between 2017 and 2023 (*n*=111; REC London – South East 16/LO/2136; Strange et al., 2026). Peripheral blood was collected in serum separator tubes, allowed to clot for 30 minutes, centrifuged at 2350 x *g* for 10 minutes at 4°C, aliquoted, frozen overnight at -80°C and subsequently stored in VPN. Samples had not previously undergone freeze-thaw cycles and were analysed using Olink Explore HT at CHG in 2026. Samples from patients with ALS, individuals with other neurologic conditions and healthy volunteers were balanced over two plates, and NPX were intensity normalised. The primary survival endpoint was death. These data were used for both significant serum protein Cox replication and external validation of multivariable Cox models.

### Statistical analysis

Statistical analysis was performed in R (v4.4.0). No *a priori* power calculations were performed. Using the first (baseline) sample from each patient, individual CSF and serum proteins were analysed using Cox proportional hazards models, with time from sampling to death or censoring as the outcome. Models were adjusted for age at sampling and sex, and stratified by sample collection cohort using package ‘survival’. *P*-values were corrected for multiple testing using the Benjamini-Hochberg false discovery rate (FDR) procedure, with FDR-adjusted p-values<0.05 considered statistically significant (Benjamini et al., 1995). Wald statistics were extracted to compare effect estimates for proteins measured in both CSF and serum.

### Functional enrichment analysis

Gene set variation analysis (GSVA) was performed in the discovery Oxford PEA data for CSF and serum separately, with Gene Ontology biological process, cellular component and molecular function, Kyoto Encyclopaedia of Genes and Genomes (KEGG) and Reactome pathways. Analyses used packages ‘GSVA’, ‘clusterProfiler’ and ‘msigdbr’, with gene sets restricted to 15-500 genes. Associations between pathway scores and survival were assessed using Cox models adjusted for age at sampling and sex, stratified by sample collection cohort, and with *p*-values adjusted by FDR procedure.

### Lasso Cox regression

To identify PEA proteins that improved prognostic prediction beyond established clinical predictors, repeated nested cross-validation with least absolute shrinkage and selection operator (lasso) Cox regression was performed separately for CSF and serum using packages ‘glmnet’, ‘nestedcv’ and ‘missForest’. Lasso applies an L1 penalty, proportional to the sum of the absolute regression coefficients, to the model loss function, shrinking some coefficients to zero and thereby excluding predictors (Tibshirani, 1996). Nested cross-validation enables hyperparameter tuning, feature selection and out-of-sample performance estimation within a dataset by optimising the regularisation parameter using inner loops, and evaluating prediction performance using outer loops (Iizuka et al., 2003; Varma et al., 2006). Repeating this process enables assessment of feature selection and prediction performance stability.

Potential ENCALS clinical predictors (age at symptom onset, bulbar onset, *C9orf72* hexanucleotide repeat expansion (HRE), definite El Escorial status, diagnostic delay, frontotemporal dementia, FVC and progression rate) were included if missingness was <50%, alongside sex and stratification by sample collection cohort (Westeneng et al., 2018). Continuous clinical predictors were log_2_-transformed and median-centred. Within each cross-validation loop, missing data for included predictors were imputed from the other included predictors using a 50-tree random forest, and proteins in the lowest 20% of variance were excluded (Bommert et al., 2021). Ten repeats of nested cross-validation were performed, using three inner and outer folds for CSF and 10 inner and outer folds for serum. Within each inner loop, the most regularised model within one standard error of the minimum partial likelihood was selected. Proteins selected in at least seven of 10 repeats were considered selected.

Internal model performance was assessed using mean out-of-sample predictions across the repeated nested cross-validation. Final multivariable coefficients were estimated using post-lasso unpenalised Cox regression, with 50 imputations by chained equations (predictive mean matching) using package ‘mice’ (Rubin, 1987). For external validation, final Oxford model coefficients were frozen and applied to the Sheffield serum PEA validation cohort to generate linear predictors. Clinical variables in the Sheffield cohort were centred and scaled to make estimates comparable with the discovery dataset. Oxford and Sheffield PEA data were generated by independent experiments and so while similarly log_2_-transformed were not explicitly centre-matched.

Discrimination was assessed using Harrell’s concordance (*c*)-statistic and inverse probability of censoring weighting time-dependent area under the receiver operating characteristic curve (AUC) at 1-5 years post-sampling using packages ‘survival’, ‘riskRegression’ and ‘timeROC’. Calibration was assessed using the calibration slope, estimated by fitting a Cox model with linear predictor as the sole covariate, and by comparing predicted mean survival with Kaplan-Meier survival curves across four risk groups defined by the 16^th^, 50^th^ and 84^th^ centiles in the linear predictors (corresponding approximately to the mean ± SD of a normal distribution; Royston et al., 2013). Absolute survival predictions in the external validation data were calculated using the Breslow-estimated baseline hazard from the AMBRoSIA development cohort, reflecting shared pre-analytic procedures. Finally, selected proteins were assessed for correlations with clinical variables and with each other in the Oxford data using pairwise-complete Spearman rank correlations adjusted for sample collection cohort.

### Longitudinal analysis

Linear mixed-effects models (LMMs) were fitted separately for each CSF and serum protein using package ‘nlme’ in the Oxford PEA data. Models included patient-specific random intercepts and slopes, with time from first sampling, age, sex and sample collection cohort as fixed-effects. *P*-values for time-associated effects were adjusted using the Benjamini-Hochberg procedure, and Wald statistics were extracted to compare proteins measured in both CSF and serum.

### Bayesian joint modelling

Proteins selected by lasso Cox regression and significantly associated with time in the LMM analyses were evaluated using event-time plots to identify candidates for exploratory joint modelling. Bayesian joint models were fitted using package ‘JMbayes2’, integrating LMM and Cox submodels. The LMM submodel included protein abundance as the outcome, linear time, patient-specific random intercepts and slopes, and age, sex and sample collection cohort as covariates. The Cox submodel included age, sex and samples collection cohort as covariates. Functional forms for both current protein value and its rate of change (slope) were specified. Markov chain Monte Carlo (MCMC) settings were progressively increased until convergence was achieved, defined by Gelman-Rubin Rhat<1.10 and satisfactory diagnostic plots.

### Mendelian randomisation

Two-sample Mendelian randomisation (MR) was used to to evaluate genetic associations between plasma protein abundance and ALS survival. Protein GWAS data were obtained from the UK Biobank Pharma Proteomics Project (UKB-PPP), comprising 2,940 targets (2,923 unique proteins) measured using Olink Explore 3072 in 54,219 European participants (Sun et al., 2023). ALS survival summary statistics were obtained from a GWAS of 4,256 patients with ALS, of whom 3,125 (73.4%) had died (median survival 32.8 months) (Fogh et al., 2016). ALS risk summary statistics were obtained from Project MinE (27,205 cases and 110,881 European-ancestry controls) (van Rheenen et al., 2021).

Genetic instruments were cis-acting biallelic single-nucleotide polymorphisms (SNPs) within 1 Mb of the encoding gene (cis-pQTLs), with minor allele frequency (MAF)>0.01, genome-wide significance (p-value<5E-08), linkage disequilibrium (LD) independence (R²<0.001, based on 10,000 unrelated European UK Biobank participants), and F-statistic≥10 (Lawlor et al., 2008). MR estimates were obtained using random-effects inverse-variance weighted (IVW) or Wald ratio methods, as appropriate, using package ‘TwoSampleMR’ (Bowden et al., 2019). P-values were adjusted using the FDR procedure.

For nominally associated proteins with multiple instruments, weighted median estimates and MR-Egger intercepts were used as sensitivity analyses to assess robustness to invalid instruments and directional pleiotropy, respectively (Bowden et al., 2016; Bowden et al., 2015). Colocalisation of protein and survival association signals across the cis region was assessed using package ‘coloc’ with default priors; PP.H4>0.8 was considered evidence of colocalisation (Zuber et al., 2022; Giambartolomei et al., 2014).

### Rare variant analysis

Rare variant burden testing was performed using Project MinE data freeze 2 whole-genome sequencing (WGS) data: 5,954 ALS cases and 2,238 controls; 5,247 had survival data, including 4,345 deaths (Project MinE ALS Sequencing Consortium, 2018). Variants were annotated using the Ensembl Variant Effect Predictor and analysed using package ‘RVAT’ (McLaren et al., 2010). Missense variants were defined as those annotated as missense, splice region, or stop variants, with a Combined Annotation-Dependent Depletion (CADD) score≥15 and MAF< 0.001 in the Genome Aggregation Database (gnomAD) (Kircher et al., 2014). Loss-of-function (LOF) variants were defined as splice, stop-gained or frameshift variants with SpliceAI scores> 0.8 and MAF< 0.001 in gnomAD.

Case-control associations were tested using SKAT-O and Firth logistic regression, adjusted for sex, sequencing platform, and the first 10 PCs (Jaganathan et al., 2019; Lee et al., 2012; Firth, 1993). Survival associated were evaluated using Cox regression of missense and LOF burden scores, adjusted for age at symptom onset, sex, sequencing platform, and the first 20 PCs.

## RESULTS

Following QC, proteomic data comprised 1,095 samples from 426 people with ALS: 244 CSF samples from 139 participants, and 851 serum samples from 392 participants. Paired first-visit CSF and serum samples were available for 104 participants. A total of 2,884 proteins were quantified in CSF using the Olink Explore 3072 platform, and 5,416 proteins in serum using Olink Explore HT. Baseline characteristics are summarised in **Table 1**, with cohort-specific stratifications provided in **Supp. Tables 2 and 3**.

**Table 1:** Clinical characteristics of patients with ALS at first CSF and serum sampling, with longitudinal sample profiles. Non-normally distributed continuous variables are presented as median (interquartile range [Q1-Q3]); normally distributed continuous variables are presented as mean (standard deviation [SD]); and categorical variables are presented as count (*n*) and percentages (%). Median survival from symptom onset calculated using the Kaplan-Meier method. Missing data are indicated where applicable. Abbreviations: ECAS – Edinburgh Cognitive and Behavioural ALS Screen; LMN – lower motor neuron; UMN – upper motor neuron; ΔALSFRS-R/ month – estimated monthly change in Revised ALS Functional Rating Scale from symptom onset to sampling.

|  | CSF<br>(*n*=139) | Serum<br>(*n*=392) |
|---|---|---|
| Sex (*n*, %) |  |  |
| Female | 40 (28.8%) | 127 (32.4%) |
| Male | 99 (71.2%) | 265 (67.6%) |
| Site of onset (*n*, %) |  |  |
| Bulbar | 34 (24.5%) | 95 (24.2%) |
| Non-bulbar | 105 (75.5%) | 297 (75.8%) |
| Age at onset (years) |  |  |
| Median (Q1-Q3) | 58.7 (51.0-67.4) | 60.9 (51.5-68.5) |
| Diagnostic delay (years) |  |  |
| Median (Q1-Q3) | 1.03 (0.65-1.70) | 1.07 (0.65-1.79) |
| Missing (%) | 41 (29.5%) | 96 (24.5%) |
| Age at sampling (years) |  |  |
| Median (Q1-Q3) | 61.7 (53.5-69.4) | 62.8 (54.1-70.5) |
| Onset to sampling time (years) |  |  |
| Median (Q1-Q3) | 1.61 (1.10-2.45) | 1.57 (0.94-2.67) |
| Diagnosis to sampling time (years) |  |  |
| Median (Q1-Q3) | 0.47 (0.04-0.97) | 0.38 (0.00-0.79) |
| Missing (%) | 41 (29.5%) | 96 (24.5%) |
| Pathogenic variant (*n*, %) |  |  |
| *C9orf72* | 17 (12.2%) | 33 (8.4%) |
| *SOD1* | 2 (1.4%) | 4 (1.0%) |
| Other | 2 (1.4%) | 7 (1.8%) |
| Negative | 11 (7.9%) | 51 (13.0%) |
| Not tested | 107 (77.0%) | 297 (75.8%) |
| ECAS (total score) |  |  |
| Median (Q1-Q3) | 113 (103-121) | 115 (106-120) |
| Missing (%) | 66 (47.5%) | 274 (69.9%) |
| Forced vital capacity (%, predicted) |  |  |
| Mean (SD) | 81 (24) | 80 (21) |
| Missing (%) | 91 (65.5%) | 347 (88.5%) |
| LMN-UMN burden score |  |  |
| Mean (SD) | -0.8 (4.2) | -1.1 (4.8) |
| Missing (%) | 88 (63.3%) | 291 (74.2%) |
| Progression rate (ΔALSFRS-R/ month) |  |  |
| Median (Q1-Q3) | 0.45 (0.28-0.71) | 0.45 (0.26-0.74) |
| Missing (%) | 11 (7.9%) | 146 (37.2%) |
| Outcome (*n*, %) |  |  |
| Dead |  |  |
| Censored | 88 (63.3%) | 263 (67.1%) |
| Median survival from onset (years, 95% CI) | 51 (36.7%)<br>4.3 (3.6-6.1) | 129 (32.9%)<br>3.9 (3.6-4.6) |
| Median survival from first sampling (years, 95% CI) | 2.1 (1.9-2.6) | 2.0 (1.8-2.2) |
| Samples by visit number (*n*) |  |  |
| 1 | 139 (57.0%) | 392 (46.1%) |
| 2 | 66 (27.0%) | 229 (26.9%) |
| 3 | 26 (10.7%) | 125 (14.7%) |
| 4 | 11 (4.5%) | 65 (7.6%) |
| 5 | 2 (0.8%) | 29 (3.4%) |
| 6 | - | 9 (1.1%) |
| 7 | - | 2 (0.2%) |
| Total | 244 (100%) | 851 (100%) |

Research cohorts differed in pre-analytic variables, including storage duration, temperature and freeze-thaw history. These differences were also reflected in CSF and serum PCA profiles (**Supp. Table 2-3, Supp. Fig. 4-6**). As these factors were entwined with sample collection cohort and potentially influenced proteomic profiling, collection cohort was included as a covariate in subsequent analyses. Samples from the six collection cohorts were evenly distributed across plates, and there were no significant differences in survival between plates (**Supp. Fig. 7-9**). Kaplan-Meier analyses showed no overall differences in survival between research cohorts in either CSF or serum (**Supp. Fig. 10-11**). Concordance between PEA and commercial immunoassays for NEFL measured in matched samples was high in both CSF and serum (*R*^2^ 0.53-0.94; all *p*-values<0.001) (**Supp. Fig. 12**).

### Cox regression identifies prognostic proteins and survival-associated biological pathways in CSF and serum

Cross-sectional univariable Cox models identified five CSF and 57 serum proteins associated with survival after FDR correction (**Fig. 1A-B, Supp. Table 4**-**5**). Consistent with previous studies, higher NEFL levels were associated with shorter survival in both CSF (hazard ratio [HR]=2.05 per 1 NPX increase [95% CI 1.63-2.57], *p*_adjust_=8.42E-07) and serum (HR=2.10 [1.84-2.39], 3.28E-25). Notably, CSF tropomyosin 3 (TPM3, HR=1.89 [1.56-2.31], *p*_adjust_=6.36E-07) and serum peripherin (PRPH, HR=1.90 [1.62-2.22], *p*_adjust_=2.10E-12) also strongly associated with shortened survival. Of 2,810 proteins measured in both biofluids, NEFL was the only protein associated with survival in both (**Fig. 1C**).

**Figure 1:**
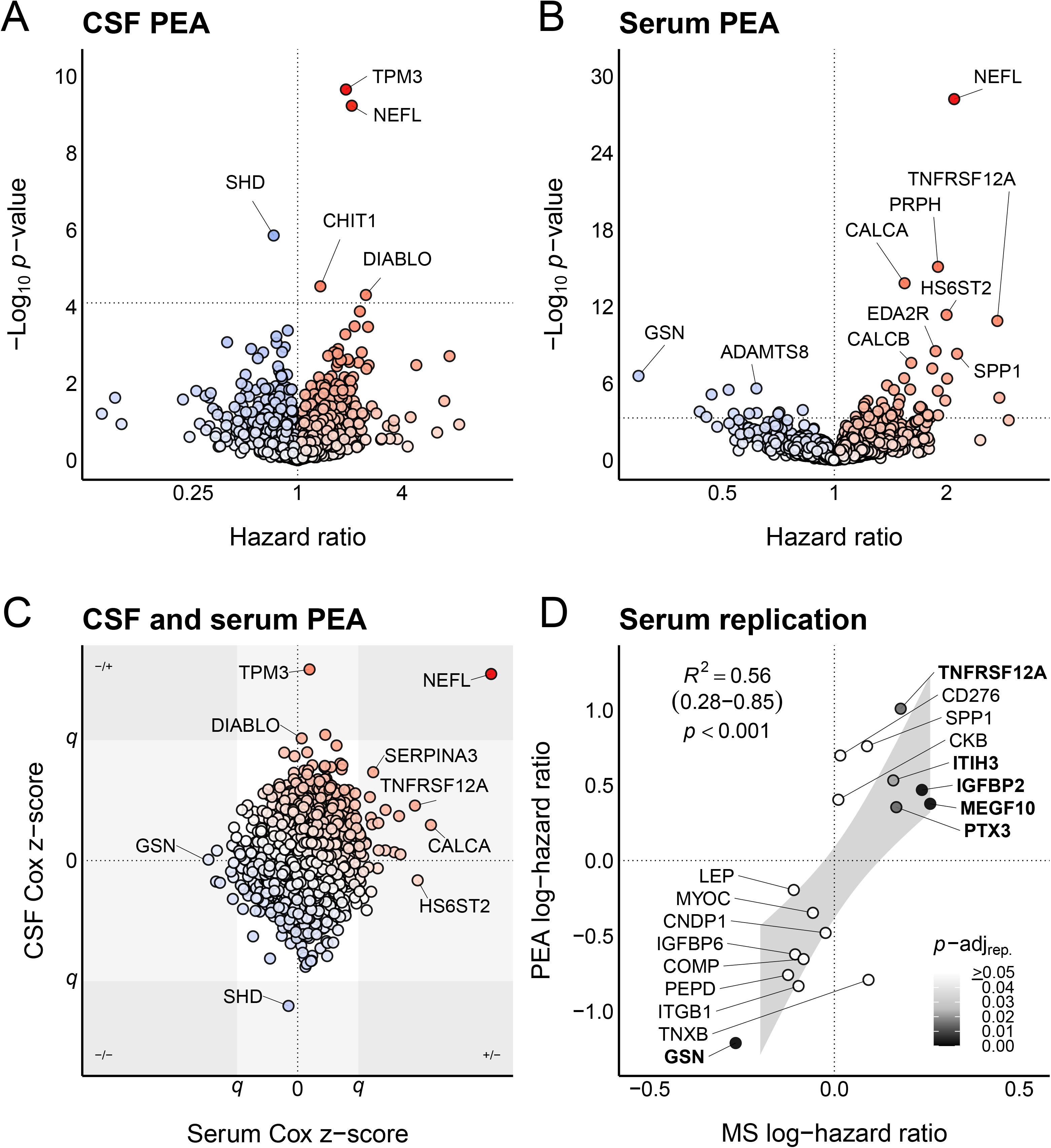
Volcano plots of hazard ratios (HRs) and *p*-values for each protein from adjusted Cox models in (**A**) CSF and (**B**) serum, with relative significance of proteins in both shown in (**C**). Dotted lines indicate the 5% false discovery rate (FDR) threshold (*q*). The colour gradient represents the Wald statistic from blue (negative) to red (positive) in (**A**) and (**B**), and a normalised composite of the CSF and serum Wald statistics in (**C**). (**D**) Discovery versus replication log-HRs for significant PEA serum proteins also detected by mass spectrometry (MS), shown with bootstrapped summary statistics from a simple linear regression with 95% confidence intervals. Proteins with a replication FDR <0.05 labelled in bold. Abbreviations: ADAMTS8 – a disintegrin and metallopeptidase with thrombospondin type 1 motif 8; CALCA – calcitonin; CALCB – calcitonin gene-related peptide 2; CD276 – CD276 antigen; CHIT1 – chitotriosidase-1; CKB – creatine kinase B-type; CNDP1 – beta-Ala-His dipeptidase; COMP – cartilage oligomeric matrix protein; DIABLO – diablo IAP-binding mitochondrial protein; EDA2R – tumor necrosis factor receptor superfamily member 27; GSN – gelsolin; HS6ST2 – heparan sulfate 6-O-sulfotransferase 2; IGFBP2 – insulin-like growth factor-binding protein 2; IGFBP6 – insulin-like growth factor-binding protein 6; ITGB1 – integrin beta-1; ITIH3 – inter-alpha-trypsin inhibitor heavy chain H3; LEP – leptin; MEGF10 – multiple epidermal growth factor-like domains protein 10; MYOC – myocilin; NEFL – neurofilament light chain; PEPD – Xaa-Pro dipeptidase; PRPH – peripherin; PTX3 – pentraxin-related protein PTX3; SERPINA3 – alpha-1-antichymotrypsin; SHD – SH2 domain-containing adapter protein D; SPP1 – osteopontin; TNFRSF12A – tumor necrosis factor receptor superfamily member 12A; TNXB – tenascin-X; TPM3 – tropomyosin alpha-3 chain.

Seventeen of the 57 candidate survival-associated serum proteins were also quantified by MS in the independent Tel Aviv cohort of 349 people with ALS (**Supp. Tables 5-6**, **Supp. Fig. 13**). Sixteen (94.1%) showed concordant directions of association with the discovery analysis (one-tailed binomial *p*-value<0.001) and sixe (35.3%) replicated after FDR correction. Effect estimates were also strongly correlated between PEA and MS (*R*^2^=0.56 [95% CI 0.28-0.85], *p*-value<0.001; **Fig. 1D**).

In the independent Sheffield serum PEA replication of 111 people with ALS, 52 of 57 proteins (91.2%) showed concordant directions of association (*p*-value=3.20E-11), with 24 (42.1%) remaining significant after FDR correction (**Supp. Tables 5-6**). Effect estimates were again strongly correlated (*R*^2^=0.44 [95% CI 0.19-0.68], *p*-value=1.82E-08). Three serum proteins were significantly associated with survival across all three Cox analyses: gelsolin (GSN), inter-alpha-trypsin inhibitor heavy chain H3 (ITIH3) and tumor necrosis factor receptor superfamily member 12A (TNFRSF12A).

Functional enrichment analysis using first-visit Oxford CSF and serum PEA data identified 159 pathways associated with survival, including terms relating to interleukin signalling, extracellular matrix organisation, nervous system and muscle development (**Supp. Tables 7-11**). No pathways were significantly associated with survival in both compartments.

### Lasso Cox identifies an eight-serum protein panel that enhances clinical predictors

Five established clinical predictors were available for more than 50% of first-visit CSF and serum PEA samples and were therefore included in models along with sex and stratification by sample collection cohort: age at symptom onset (no missing data), bulbar vs non-bulbar onset (no missing data), diagnostic delay (29.5% missing in CSF; 24.5% missing in serum), progression rate (7.9% missing in CSF; 37.2% missing in serum) and *C9orf72* HRE (12.2% and 8.4% HRE-positive in CSF and serum, respectively). In the repeated nested cross-validation lasso Cox analysis, no CSF PEA proteins were selected in any of the 10 repeats to augment these clinical predictors. The clinical predictor-only model in the CSF cohort achieved a mean (SD) out-of-sample *c*-statistic of 0.70 (0.02).

In the serum lasso analysis, eight proteins were selected in at least seven of the 10 repeats: a disintegrin-like and metalloprotease with thrombospondin motifs 8 (ADAMTS8, *n*=10), ADP-ribosyltransferase 3 (ART3, *n*=10), CALCA (annotated to calcitonin, *n*=10), tumor necrosis factor receptor superfamily member 27 (EDA2R, *n*=10), heparan sulfate 6-O-sulfotransferase 2 (HS6ST2, *n*=9), myosin light chain 11 (MYL11, *n*=7), NEFL (*n*=10) and peripherin (PRPH, *n*=9; **Table 2**, **Fig. 2A-C**). Raw count density plots for these proteins are shown in **Supp. Fig. 15**. Four additional proteins were selected in fewer than seven repeats and therefore not retained: dapper homolog 1 (DACT1, *n*=1), growth/differentiation factor 8 (MSTN, *n*=4), thialysine N-epsilon-acetyltransferase (SAT2, *n*=6) and histone-lysine N-methyltransferase SETD5 (SETD5, *n*=6). Of the eight selected proteins, only ADAMTS8 and ART3 were positively associated with survival.

**Table 2:** Comparison of three baseline multivariable Cox proportional hazards models: clinical predictors only, clinical predictors and serum NEFL, and clinical predictors and lasso-selected serum proteins. Results are presented as hazard ratios (HRs) with 95% confidence intervals (Cis). Numeric clinical predictors are scaled per one standard deviation, and protein predictors are reported per NPX. Abbreviations: ADAMTS8 – a disintegrin and metallopeptidase with thrombospondin type 1 motif 8; ART3 – ecto-ADP-ribosyltransferase 3; AUC – area under the curve; *c*-statistic – concordance statistic; CALCA – calcitonin; CI – confidence interval; EDA2R – tumor necrosis factor receptor superfamily member 27; HR – hazard ratio; HS6ST2 – heparan sulfate 6-O-sulfotransferase 2; lasso – least absolute shrinkage and selection operator; MYL11 – myosin regulatory light chain 11; PRPH – peripherin.

|  | Clinical variables only | Clinical variables and serum NEFL | Clinical variables and lasso serum |
|---|---|---|---|
| Clinical variables |  |  |  |
| Age at onset | 1.38 (1.20-1.60) | 1.34 (1.16-1.54) | 1.36 (1.12-1.65) |
| Bulbar onset | 1.62 (1.17-2.24) | 1.08 (0.79-1.48) | 1.72 (1.20-2.47) |
| Diagnostic delay | 0.92 (0.76-1.10) | 1.03 (0.86-1.23) | 0.96 (0.80-1.16) |
| Male sex | 1.22 (0.90-1.65) | 1.24 (0.93-1.67) | 1.14 (0.82-1.61) |
| Progression rate | 1.33 (1.04-1.70) | 1.18 (0.94-1.49) | 1.12 (0.90-1.40) |
| ***C9orf72*** HRE | 1.18 (0.79-1.76) | 1.10 (0.73-1.64) | 1.08 (0.71-1.65) |
| Serum proteins |  |  |  |
| ADAMTS8 | - | - | 0.71 (0.55-0.91) |
| ART3 | - | - | 0.51 (0.39-0.67) |
| CALCA | - | - | 1.19 (1.02-1.39) |
| EDA2R | - | - | 1.11 (0.82-1.49) |
| HS6ST2 | - | - | 1.03 (0.78-1.36) |
| MYL11 | - | - | 1.10 (0.97-1.25) |
| NEFL | - | 1.97 (1.70-2.28) | 2.03 (1.69-2.42) |
| PRPH | - | - | 1.41 (1.14-1.74) |
| Internal validation |  |  |  |
| *c*-statistic (SD) | 0.66 (0.01) | 0.74 (0.01) | 0.74 (0.01) |
| Calibration slope (SD) | 0.85 (0.05) | 0.85 (0.02) | 1.06 (0.05) |
| External validation |  |  |  |
| *c*-statistic (95% CI) | 0.69 (0.62-0.75) | 0.74 (0.69-0.79) | 0.78 (0.73-0.82) |
| 1-year AUC (95% CI) | 0.75 (0.64-0.86) | 0.84 (0.76-0.92) | 0.89 (0.83-0.96) |
| 2-year AUC (95% CI) | 0.76 (0.67-0.85) | 0.80 (0.72-0.88) | 0.85 (0.78-0.92) |
| 3-year AUC (95% CI) | 0.81 (0.73-0.89) | 0.86 (0.79-0.93) | 0.88 (0.82-0.95) |
| 4-year AUC (95% CI) | 0.81 (0.73-0.89) | 0.87 (0.79-0.94) | 0.88 (0.81-0.95) |
| 5-year AUC (95% CI) | 0.88 (0.81-0.96) | 0.84 (0.74-0.93) | 0.89 (0.82-0.97) |
| Calibration slope (95% CI) | 0.99 (0.71-1.27) | 0.92 (0.69-1.16) | 0.75 (0.58-0.93) |

**Figure 2:**
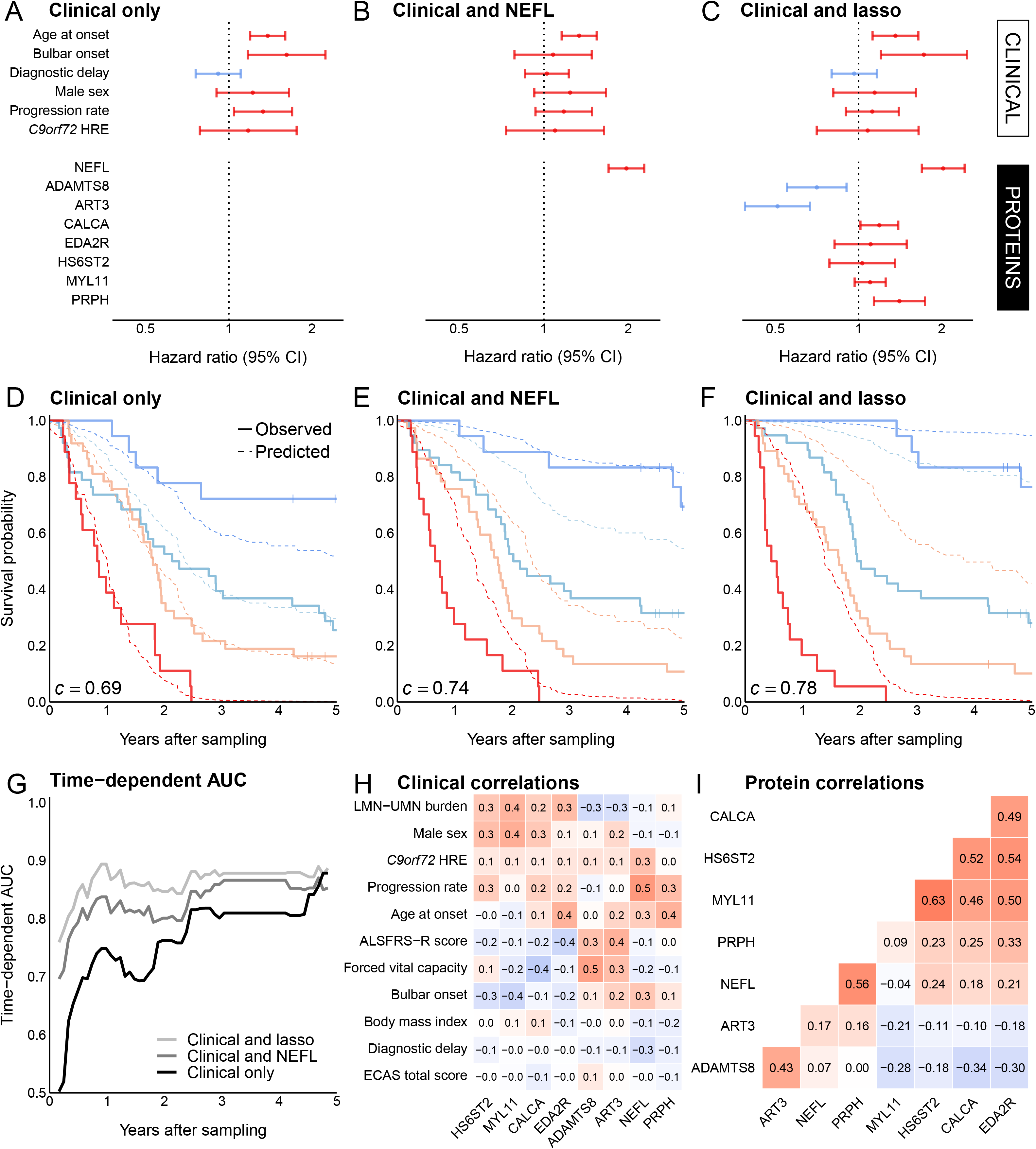
Summary of multivariable Cox proportional hazards models incorporating (**A**) clinical predictors only, (**B**) clinical predictors and serum NEFL, and (**C**) clinical predictors and lasso-selected serum proteins, fitted using post-lasso unpenalised Cox regression in Oxford cohort data. Red indicates variables with hazard ratio >1, blue <1. (**D**-**F**) External validation discrimination and calibration performance of these Cox models shown in Sheffield PEA data by comparing predicted survival with observed Kaplan-Meier curves across risk groups defined by 16^th^, 50^th^ and 84^th^ centiles of predicted survival (lowest risk - blue; highest - red), shown with concordance (*c*)-statistic. (**G**) Comparison of time-dependent AUC profiles of these three models trained in Oxford PEA data and externally validated in Sheffield data. Spearman’s rank correlation coefficients between these lasso-selected serum proteins and (**H**) pairwise-complete clinical features and (**I**) each other, with rows and columns hierarchically clustered. Abbreviations: ADAMTS8 – a disintegrin and metallopeptidase with thrombospondin type 1 motif 8; ALSFRS-R – Revised ALS Functional Rating Scale; ART3 – ecto-ADP-ribosyltransferase 3; *C9orf72* – C9orf72-SMCR8 complex subunit; CALCA – calcitonin; ECAS – Edinburgh Cognitive and Behavioural ALS Screen; EDA2R – tumor necrosis factor receptor superfamily member 27; HRE – hexanucleotide repeat expansion; HS6ST2 – heparan sulfate 6-O-sulfotransferase 2; MYL11 – myosin regulatory light chain 11; NEFL – neurofilament light chain; PRPH – peripherin.

Internal evaluation using out-of-sample predictions from the repeated nested cross-validation showed that the clinical predictors and serum lasso model achieved a mean (SD) out-of-sample *c*-statistic of 0.74 (0.01), compared with 0.66 (0.01) for clinical predictors alone and 0.74 (0.01) for clinical predictors plus serum NEFL (**Table 2**). Internal calibration slopes were closest to 1 in the lasso model, with mean (SD) values of 1.06 (0.05) for the clinical predictors and lasso-selected proteins, compared with 0.85 (0.05) for the clinical predictors alone and 0.85 (0.02) for clinical predictors plus serum NEFL (**Table 2**).

External validation of the Cox models developed in the Oxford serum cohort was performed in the independent Sheffield serum cohort (**Supp. Table 6**; **Supp. Fig. 14**; **Fig. 2D-G**). Discrimination progressively improved across the three models: the *c*-statistic increased from 0.69 (95% CI 0.62-0.75) for clinical predictors alone to 0.74 (0.69-0.79) with the addition of NEFL, and further to 0.78 (0.73-0.82) with the addition of lasso-selected proteins (**Table 2**; **Fig. 2D-F**). Both pairwise increases were borderline significant: addition of NEFL, Δ *c*-statistic 95% CI 0.00-0.11, *p*-value=0.051; and addition of lasso-selected proteins beyond NEFL, Δ 0.00-0.08, *p*-value=0.066. Similarly, time-dependent AUCs from 1 to 5 years were highest for the model incorporating clinical variables and lasso-selected proteins. The improvement over the NEFL model was statistically significant only at 1 year (AUC 0.89 versus 0.84, Δ 95% CI 0.00-0.11, *p*-value=0.047; **Table 2**; **Fig. 2G**).

Calibration plots comparing predicted and observed survival in the external data indicated increasing underestimation of risk as additional protein predictors were incorporated (**Fig. 2D-F**, **Table 2**). This was reflected in the calibration slopes, which progressively decreased from 0.99 (0.71-1.27) for clinical predictors alone to 0.92 (0.69-1.16) with the addition of NEFL, and 0.75 (0.58-0.93) with lasso-selected proteins (**Table 2**).

### Clinical and biochemical correlations of lasso-selected serum proteins reveal groupings and peripheral compartment specificity

Correlation analysis of the eight lasso-selected serum proteins and clinical variables revealed distinct patterns of association (**Fig. 2H-I**). NEFL and peripherin were strongly positively correlated with each other (ρ=0.56, *p*-value=8.15E-33), and both were positively associated with progression rate (NEFL ρ=0.51, *p*-value=2.03E-17; peripherin ρ=0.34, *p*-value=6.06E-08) and age at symptom onset (NEFL ρ=0.25, *p*-value=5.27E-07; peripherin ρ=0.39, *p*-value=2.65E-15). Similarly, ADAMTS8 and ART3 were positively correlated with each other (ρ=0.43, *p*-value=4.32E-19), FVC (ADAMTS8 ρ=0.47, *p*-value=2.12E-03; ART3 ρ=0.33, *p*-value=3.66E-02) and ALSFRS-R score (ADAMTS8 ρ=0.32, *p*-value=5.29E-07; ART3 ρ=0.39, *p*-value=2.11E-10).

CALCA, EDA2R, HS6ST2 and MYL11 were strongly positively correlated with one another across all pairware comparisons (ρ=0.46-0.63, all *p*<8.00E-22). All four proteins were positively associated with LMN-UMN burden score (ρ=0.24-0.38) and male sex (ρ=0.07-0.41), and negatively with bulbar onset (ρ=-0.08 to -0.37). However, associations between CALCA and LMN-UMN burden score or bulbar onset, and between EDA2R and sex, were not statistically significant. EDA2R was also negatively associated with ALSFRS-R score (ρ=-0.36, *p*-value=5.24E-09) and positively associated with age (ρ=0.39, *p*-value=1.93E-15). Six of the eight serum proteins (ADAMTS8, ART3, CALCA, EDA2R, HS6ST2 and NEFL) were also measured in CSF. In 104 paired CSF and serum samples, NEFL was the only protein that showed a significant correlation between compartments (**Supp. Fig. 16**).

### Longitudinal analysis reveals dynamic protein alterations in CSF and serum

Longitudinal analyis identified significant changes in three CSF proteins, all of which increased over time, and 324 serum proteins, of which 273 increased and 51 decreased (**Fig. 3A-B, Supp. Table 4-5**). Leucine-rich alpha-2-glycoprotein (LRG1) was the only protein showing significant longitudinal changes in both compartments, increasing by 0.024 NPX/month in CSF (95% CI 0.014 to 0.035, *p*_adjust_=0.019) and 0.010 NPX/month in serum (0.006 to 0.014, *p*_adjust_=1.90E-04; **Fig. 3C**).

**Figure 3:**
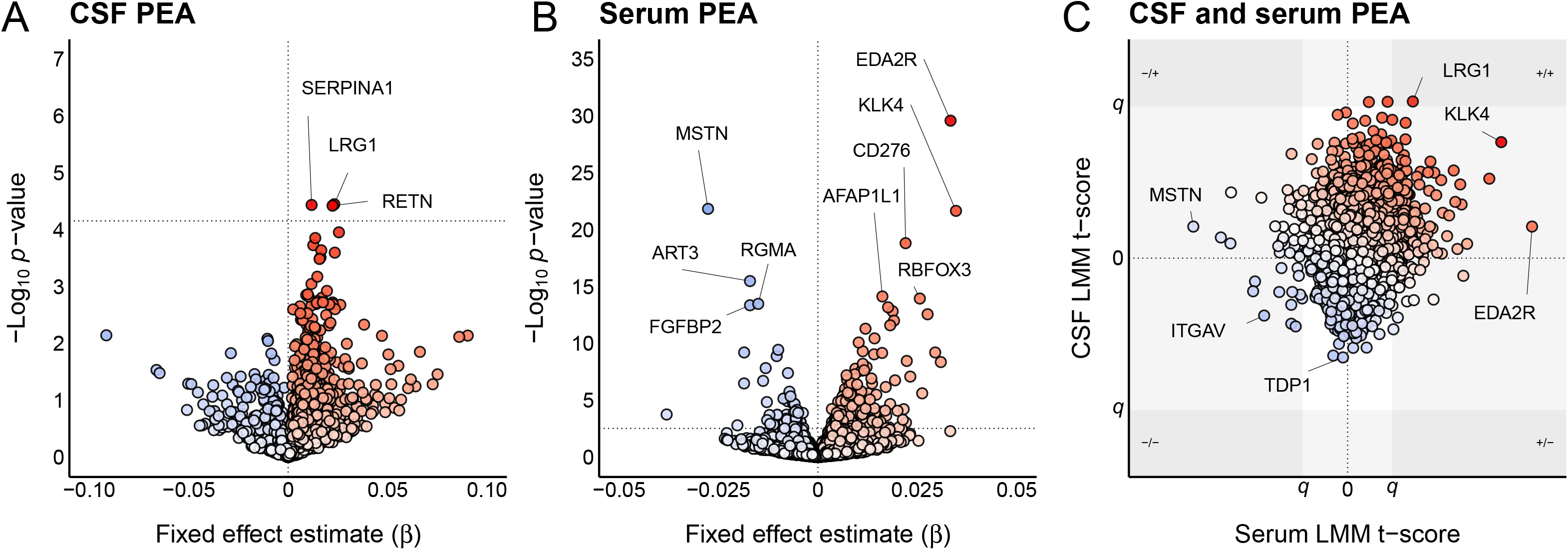
Volcano plots showing linear mixed-effects model (LMM) fixed effect estimates (β; monthly change in NPX) and *p*-values for each protein in (**A**) CSF and (**B**) serum, with relative significance of proteins measured in both matrices shown in (**C**). Abbreviations: AFAP1L1 – actin filament-associated protein 1-like 1; ART3 – ecto-ADP-ribosyltransferase 3; CD276 – CD276 antigen; EDA2R – tumor necrosis factor receptor superfamily member 27; FGFBP2 – fibroblast growth factor-binding protein 2; ITGAV – integrin alpha-V; KLK4 – kallikrein-4; LRG1 – leucine-rich alpha-2-glycoprotein; MSTN – growth/differentiation factor 8; RBFOX3 – RNA binding protein fox-1 homolog 3; RETN – resistin; RGMA – repulsive guidance molecule A; SERPINA1 – alpha-1-antitrypsin; TDP1 – tyrosyl-DNA phosphodiesterase 1.

Six of the eight lasso-selected serum proteins also showed significant longitudinal changes. ADAMTS8 and ART3 decreased over time, with estimated changes of - 0.013 NPX/month (-0.019 to -0.007, *p*_adjust_=7.13E-04) and -0.016 NPX/month (-0.020 to -0.012, *p*_adjust_=2.74E-12), respectively. Conversely, CALCA, EDA2R, HS6ST2 and MYL11 increased over time, with estimated changes of 0.028 (0.021 to 0.035, *p*_adjust_=3.70E-11), 0.035 (0.030 to 0.040, *p*_adjust_=2.54E-29), 0.016 (0.011 to 0.021, *p*_adjust_=1.57E-07) and 0.029 NPX/month (0.020 to 0.038, *p*_adjust_=1.93E-07), respectively. NEFL and peripherin showed nominal increases over time, but these did not remain significant after FDR correction (**Supp. Table 5**).

### Bayesian joint modelling identifies survival-associated trajectories of serum CALCA, EDA2R and NEFL and potentially enables dynamic risk prediction

Six serum proteins (ADAMTS8, ART3, CALCA, EDA2R, HS6ST2 and MYL11) were selected by the lasso Cox analysis and also showed significant longitudinal associations in the LMM analysis. Their trajectories were therefore examined using event-time plots, alongside serum NEFL and ALSFRS-R as reference measures (**Fig. 4A-H**). CALCA and EDA2R showed the clearest visual associations between longitudinal trajectories and time to death or censoring and were therefore selected for joint modelling alongside NEFL. CSF proteins were not included in joint modelling due to limited longitudinal data.

**Figure 4:**
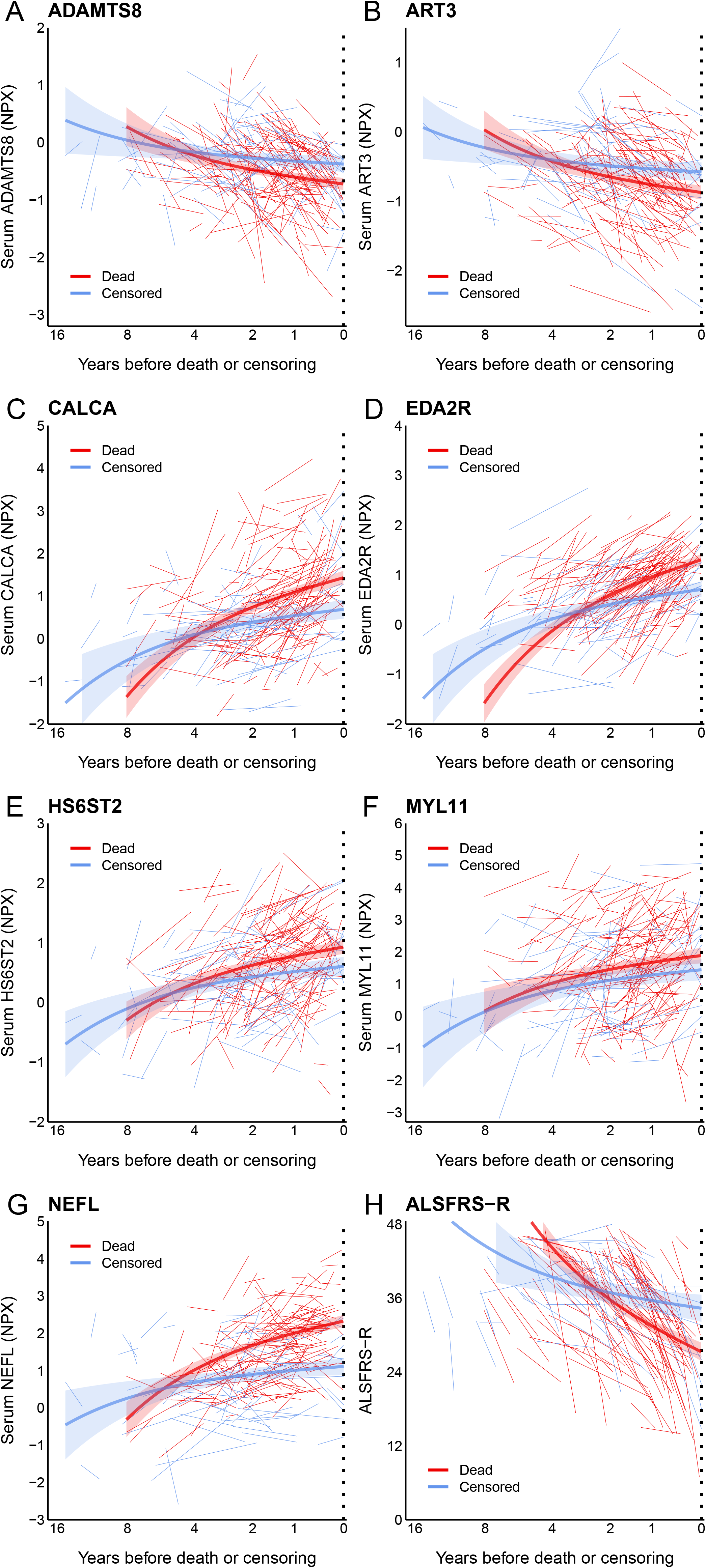
Event-time plots of longitudinal serum values in patients with ALS preceding death or censoring for (**A**) ADAMTS8, (**B**) ART3, (**C**) CALCA, (**D**) EDA2R, (**E**) HS6ST2, (**F**) MYL11, (**G**) NEFL and (**H**) ALSFRS-R. Individual lines of best fit are shown for each patient, together with group-level lines of best fit for the death and censoring groups and their 95% confidence intervals. Group-level estimates were obtained using linear mixed-effects models with an interaction between group and event time. Abbreviations: ADAMTS8 – a disintegrin and metallopeptidase with thrombospondin type 1 motif 8; ALSFRS-R – Revised ALS Functional Rating Scale; ART3 – ecto-ADP-ribosyltransferase 3; CALCA – calcitonin; EDA2R – tumor necrosis factor receptor superfamily member 27; HS6ST2 – heparan sulfate 6-O-sulfotransferase 2; MYL11 – myosin regulatory light chain 11; NEFL – neurofilament light chain.

Joint models showed that serum CALCA, EDA2R and NEFL all increased over time (**Supp. Tables 12-14**). Estimated mean monthly increases were 0.038 NPX for CALCA (95% credible interval [CrI] 0.030-0.046), 0.044 NPX for EDA2R (0.038-0.049), and 0.013 NPX for NEFL (0.006-0.020). Faster increases in all three proteins were associated with shorter survival. For each additional 0.01 NPX/month in the rate of increase, the hazard of death increased by 33% for CALCA (95% CrI 12– 72%), 67% for EDA2R (31–144%), and 42% for NEFL (0–201%). Corresponding Rhat values ranged from 1.00 to 1.05, and model diagnostic plots indicated satisfactory convergence and sampling efficiency in all three joint models (**Supp. Fig. 17-19**). These findings identify these serum proteins as candidate dynamic biomarkers, with serial measurements potentially enabling quantification of changes in individual patients’ risk over time.

### Genetic analysis of survival-associated proteins

Exploratory genetic analyses, incorporating two-sample MR, missense and LOF rare variant analyses, were integrated to investigate potential causal associations between ALS survival or risk for four of five CSF proteins, and 50 of 57 serum proteins identified as survival-associated in the adjusted univariable Cox analyses. Of the eight serum lasso proteins, genetic analyses were not possible for ART3, EDA2R, HS6ST2 or NEFL. Among proteins with available genetic data, only MMP13 showed evidence of association with survival in rare variant analysis after correction for multiple testing (β_LOF_=2.04, *p_adjust_*=0.009; **Supp. Table 15**). The direction of effect was concordant with the observational association but not with the MR estimate.

Four additional proteins showed nominally significant associations in at least one genetic analysis, with effect directions concordant with their observational findings. IFT43 (β_LOF+miss_=0.390, *p*-value=0.013) and ACP5 (β_LOF+miss_=0.380, *p*-value=0.043) were associated with survival in the combined LOF and missense rare variant analysis, while KLK4 (β_MR_=0.31, *p*-value=0.003) and SERPINA3 (β_MR_=0.15, p-value=0.048) showed nominal associations in the MR analysis. MR sensitivity analyses were possible only for KLK4; the weighted median estimate was consistent with the IVW estimate, with no evidence of directional pleiotropy (**Supp. Table 15**). Colocalisation analysis provided additional evidence of shared genetic signals between ALS survival and both KLK4 and SERPINA3.

## DISCUSSION

High-depth CSF and serum PEA in a large longitudinal ALS cohort identified five CSF and 57 serum proteins associated with survival, and an eight-protein serum panel that complements clinical parameters and NEFL in prognostic stratification. This panel improved discrimination, particularly over shorter time horizons, and captured a broader range of biological signals than NEFL alone, although its suboptimal calibration highlights the need for absolutely quantitative assay development prior to clinical translation. Longitudinal analyses identified assay trajectories associated with survival, including dynamic changes in serum CALCA and EDA2R that were associated with subsequent mortality in joint models. Exploratory genetic epidemiological analysis provided little evidence for a causal role of these protein alterations in determining survival, suggesting that many of the observed changes may be downstream consequences rather than drivers of disease progression.

The serum and CSF proteomes provided distinct information. A broader range of longitudinal and survival-associated alterations was detected in serum than in CSF, and only serum proteomic data improved prognostic models beyond established predictors. While these differences may reflect the smaller CSF samples size and reduced statistical power, it is consistent with previous findings that plasma, but not CSF, NEFL levels improve established prognostic models (Thompson et al., 2022). More broadly, the limited overlap between compartments suggest that CSF and serum capture different aspects of ALS biology. NEFL was the only protein associated with survival in both biofluids after correction for multiple comparisons, while functional enrichment analyses also showed little overlap.

The added prognostic value of the serum proteome may seem counterintuitive given the predominantly central nervous system pathology of ALS. However, survival also reflects the systemic consequences of motor system degeneration, including respiratory dysfunction, nutritional impairment, muscle atrophy and inflammation, alongside biological factors such as ageing, and these processes are reflected in the circulating proteome. Consistent with this, the clinical correlates of the optimised prognostic panel extended beyond measures of disease aggressiveness to include proteins associated with overall disability, respiratory impairment and age. The proteins also mapped to biological pathways involving inflammatory signalling, extracellular matrix and muscle contractile machinery.

Several proteins showed particularly strong associations with survival. In univariable CSF analysis, the post-synaptic actin-binding protein TPM3 showed a stronger association with survival than NEFL. CSF TPM3 has previously been reported as elevated in ALS compared with controls using both PEA and aptamer-based proteomics, in people with FTD and pathogenic TPM3 variants have been linked with congenital myopathies (Chia et al., 2025; Martilla et al., 2014; De Houwer et al., 2026). The absence of a comparable serum association may indicate a central source of this signal, implicating neuronal rather than striated muscle function in its association with survival.

Serum EDA2R, a member of the tumor necrosis factor (TNF) receptor superfamily, was associated with survival in both univariable and multivariable feature-selected analyses. Further, longitudinal increases in EDA2R were associated with shorter survival. EDA2R has previously been reported as elevated in ALS and to increase during the years preceding symptomatic onset in ALS gene carriers (Chia et al., 2025; Ran et al., 2025). These findings suggest that EDA2R may reflect biological processes that precede symptomatic disease and continue during clinical progression. EDA2R has been proposed as a tissue-independent ubiquitous marker of ageing, with activation linked to NF-kappa B and JNK signalling and to parainflammatory responses, including sarcopaenia and insulin resistance (Barbera et al., 2025; Sinha et al., 2002; Bilgic et al., 2023; Awazawa et al., 2017).

Serum CALCA and CALCB were also strongly associated with survival in univariable analysis. CALCA was one of eight proteins selected in the lasso Cox regression, and steeper longitudinal increases in CALCA were associated with shorter survival, supporting a role in measuring disease progression as well as cross-sectional risk. Plasma CALCB has previously been reported as elevated in people with ALS compared with controls, while serum CALCA increases have been observed before the symptomatic onset in individuals at genetic risk (Imam et al., 2025; Ran et al., 2025). CALCA and CALCB are annotated to the related proteins calcitonin and calcitonin gene-related peptide 2 (β-CGRP), respectively, although precise specificity of the assays is not known and may not distinguish mature from precursor or related proteoforms. Calcitonin and calcitonin gene-related peptide 1 (α-CGRP) are produced through alternative splicing of the *CALCA* gene but have different functions, with calcitonin’s primary role in calcium homeostasis and α-CGRP having roles in vasomotor function and neuromodulation. β-CGRP, encoded by *CALCB*, is proposed to have vasoactive and anti-inflammatory roles, and both peptides can readily form fibrillar aggregates (Russo et al., 2023; Reches et al., 2002).

Serum peripherin, a peripheral nerve intermediate filament protein, was also strongly negatively associated with survival in univariable analysis and was selected by lasso Cox regression. Notably, although peripherin levels were all below the reported limit of detection of the assay, the finding was reproduced with comparable magnitude in the external PEA dataset. This result contrasts with two smaller ELISA studies of CSF, serum and plasma peripherin, which reported either non-significant or positive associations with survival (Sabbatini et al., 2021; Bombaci et al., 2025). This discrepancy may relate to cohort differences, matrix effects or, given that PEA relies on the proximity of antibody probes binding to a target in solution, could therefore reflect protein fragments, complexes or isoforms not measured by sandwich ELISA. Further work is required to determine whether these discrepant findings reflect differences in platform, matrix, cohort characteristics or sample size.

Several limitations are noted. First, there is selection bias among patients with ALS motivated and able to attend a regional referral centre and donate biofluids. Although our data capture a broad range of phenotypes, it is weighted towards individuals with more slowly progressive disease and is less diverse than the global ALS population. Samples were also typically collected later than diagnosis, limiting the extrapolation of our findings to initial clinical presentation, and the collection period spanned an evolution in diagnostic criteria without explicit harmonisation.

Missing data for some established clinical predictors, including genotype, FVC and some cognitive measures, led to the exclusion of potentially relevant factors from some analyses. This may have influenced lasso Cox regression protein selection and limits the interpretation of associations between proteins and clinical features. As genotype has been associated with CSF and circulating proteomic differences in ALS, the limited genotyping in this cohort means that confounding by genetic variation cannot be excluded for some identified proteins (Chia et al., 2025; Trautwig et al., 2025). Other potentially important prognostic factors, including riluzole use and nutritional and respiratory support, were also not systematically captured. While the discrepancy in outcome definitions between the Oxford and Sheffield cohorts (death), and Tel Aviv cohort (death or tracheostomy) may influence effect estimates, the consistency of findings supports the robustness of the identified associations.

The CSF and serum cohorts differed in some clinical characteristics, including the prevalence of ALS-causing genetic variants, potentially confounding comparisons across compartments. The relatively smaller CSF sample size with consequently reduced statistical power means that some negative findings could reflect an inability to detect significance. In addition, some promising candidate ALS survival biomarkers such as ubiquitin carboxyl hydrolase L1 (UCHL1) are not included in Olink PEA and could not be assessed (Dellar et al., 2024). A number of additional protein candidates, including post-translationally modified forms and those not analysed in this work, have shown promise in discriminating ALS cases from healthy controls with high accuracy (Kim et al, 2026); our panel cannot therefore be considered definitive. The absence of exclusion criteria for recent infection, inflammatory conditions or renal dysfunction may have introduced biological confounding of circulating protein levels. Lasso Cox regression was chosen for its embedded feature selection and relative interpretability, whereas more complex modelling approaches such as random forests or neural networks might have identified different protein subsets with different validation performance.

Although significant proteins prioritised using PEA showed high concordance with MS for proteins measured on both platforms, orthogonal validation is required to confirm the identity and prognostic associations of these candidate proteins, particularly those with high CVs or uncertain LOD profiles, noting that a high proportion of samples were below LOD in a substantial proportion of samples and that intra- and inter-plate CVs were above those accepted in clinical assays. External validation of the prognostic models was limited by the relatively small validation cohort and comparisons between PEA datasets with different median-centring.

For the genetic causal analyses, instruments derived from affinity-based proteomic platforms may reflect epitope-altering variants rather than true differences in protein abundance. Furthermore, the limited number and strength of some instruments reduced power to detect causal effects of plausible magnitude. Null findings should therefore not be interpreted as excluding causality. Although deriving exposure and outcome data from populations of European genetic ancestry reduces potential bias from mismatched LD structures, it limits generalisability to other ancestral groups. Finally, case-only survival analyses are susceptible to index-event bias, whereby conditioning on disease occurrence can induce spurious associations between genetic determinants of disease susceptibility and subsequent survival, potentially biasing estimates in either direction.

In conclusion, this study identified a serum protein signature that improves prognostic stratification when combined with clinical parameters, and captures biological processes associated with ALS progression and survival. Further development of these markers could enable dynamic, personalised risk estimates, with potential applications in clinical management, patient stratification and the design and interpretation of disease-modifying therapy trials.

## Data Availability

Oxford proteomic and clinical data may be requested via the corresponding author.

## Author contributions

DGL: conceptualisation, methodology, analysis, visualisation, data curation, drafting, reviewing and editing; PP: analysis, data curation, reviewing and editing; ED: conceptualisation, analysis, reviewing and editing; HK: analysis, data curation, reviewing and editing; CVC: analysis, editing and reviewing; MW: analysis, editing and reviewing; EA: analysis, editing and reviewing; AS: analysis, editing and reviewing; DMK: analysis, editing and reviewing; TZ: analysis, editing and reviewing; PJS: conceptulatisation, data curation, editing and reviewing; AA: analysis, editing and reviewing; VD: conceptulatisation, data curation, editing and reviewing; AM: conceptualisation, data curation, editing and reviewing; JCK: conceptualisation, supervision, editing and reviewing; IM: analysis, supervision, editing and reviewing; EH: conceptualisation, data curation, supervision, editing and reviewing; AO: conceptualisation, funding acquisition, editing and reviewing; GN: conceptualisation, funding acquisition, editing and reviewing; AT: analysis, data curation, supervising, editing and reviewing; JAT: conceptualisation, funding acquisition, editing and reviewing; KT: conceptualisation, funding acquisition, data curation, supervision, editing and reviewing; MRT: conceptualisation, funding acquisition, data curation, supervision, editing and reviewing; AGT: conceptualisation, funding acquisition, data curation, supervision, editing and reviewing.

## Acknowledgements

We are grateful to participants for donating samples used in this work. We thank Beatrice Abramovich for coordinating the establishment of the ALS patient cohort at Tel Aviv Medical Center.

## Funding

This work was supported by grants and funding from UK Research and Innovation (MR/Y001095/1), the MND Association (Lester/2450/795), the Oxford-GSK Institute of Molecular and Computational Medicine and the Israeli Science Foundation (3497/21).

## Data availability

Oxford PEA and Oxford clinical data may be requested via the corresponding author.

## Code availability

All analyses were performed using standard functions within the described software packages. Individual workflows and specifications are described in the <u>Methods</u> sections.

## Notes

### Competing Interest Statement

The authors have declared no competing interest.

### Author Declarations

London - South East REC 16/LO/2136 gave ethical approval for this work. South Central Oxford Research Ethics Committee 08/H0605/85 gave approval for this work. South Central - Hampshire B REC 14/SC/1248 gave ethical approval for this work. South Central - Oxford C REC 17/SC/0277 gave ethical approval for this work. Wales REC 20/WA/0027 gave ethical approval for this work. East Midlands - Leicester Central REC 21/EM/0251 gave ethical approval for this work.

### Summary of Updates

External validation data incorporated following peer review. Abstract, introduction, methods, results, discussion, figures and supplementaries updated.

## REFERENCES

Awazawa M, Gabel P, Tsaousidou E, Nolte H, Krüger M, Schmitz J, Ackermann PJ, Brandt C, Altmüller J, Motameny S, Wunderlich FT, Kornfeld JW, Blüher M, Brüning JC. A microRNA screen reveals that elevated hepatic ectodysplasin A expression contributes to obesity-induced insulin resistance in skeletal muscle. Nat Med. 2017 Dec;23(12):1466–1473. doi: 10.1038/nm.4420.

Barbera MC, Guarrera L, Re Cecconi AD, Cassanmagnago GA, Vallerga A, Lunardi M, Checchi F, Di Rito L, Romeo M, Mapelli SN, Schoser B, Generozov EV; Molecular Genetics Group; Jansen R, de Geus EJC, Penninx B, van Dongen J, Craparotta I, Piccirillo R, Ahmetov II, Bolis M. Increased ectodysplasin-A2-receptor EDA2R is a ubiquitous hallmark of aging and mediates parainflammatory responses. Nat Commun. 2025 Feb 23;16(1):1898. doi: 10.1038/s41467-025-56918-3.

Benatar M, Macklin EA, Malaspina A, Rogers ML, Hornstein E, Lombardi V, Renfrey D, Shepheard S, Magen I, Cohen Y, Granit V, Statland JM, Heckmann JM, Rademakers R, McHutchison CA, Petrucelli L, McMillan CT, Wuu J; CReATe Consortium PGB1 Study Investigators. Prognostic clinical and biological markers for amyotrophic lateral sclerosis disease progression: validation and implications for clinical trial design and analysis. EBioMedicine. 2024 Oct;108:105323. doi: 10.1016/j.ebiom.2024.105323.

Benatar M, Zhang L, Wang L, Granit V, Statland J, Barohn R, Swenson A, Ravits J, Jackson C, Burns TM, Trivedi J, Pioro EP, Caress J, Katz J, McCauley JL, Rademakers R, Malaspina A, Ostrow LW, Wuu J; CReATe Consortium. Validation of serum neurofilaments as prognostic and potential pharmacodynamic biomarkers for ALS. Neurology. 2020 Jul 7;95(1):e59–e69. doi: 10.1212/WNL.0000000000009559.

Benjamini Y, Hochberg Y. Controlling the False Discovery Rate: A Practical and Powerful Approach to Multiple Testing. J R Stat Soc Ser B Stat Methodol. 1995;57(1):289–300. doi:10.1111/j.2517-6161.1995.tb02031.x.

Bilgic SN, Domaniku A, Toledo B, Agca S, Weber BZC, Arabaci DH, Ozornek Z, Lause P, Thissen JP, Loumaye A, Kir S. EDA2R-NIK signalling promotes muscle atrophy linked to cancer cachexia. Nature. 2023 May;617(7962):827–834. doi: 10.1038/s41586-023-06047-y.

Bombaci A, De Marco G, Casale F, Salamone P, Marchese G, Fuda G, Calvo A, Chiò A. Peripherin: A Novel Early Diagnostic and Prognostic Plasmatic Biomarker in Amyotrophic Lateral Sclerosis. Eur J Neurol. 2025 Jun;32(6):e70241. doi: 10.1111/ene.70241.

Bommert A, Welchowski T, Schmid M, Rahnenführer J. Benchmark of filter methods for feature selection in high-dimensional gene expression survival data. Brief Bioinform. 2022 Jan 17;23(1):bbab354. doi: 10.1093/bib/bbab354.

Bowden J, Del Greco M F, Minelli C, Zhao Q, Lawlor DA, Sheehan NA, Thompson J, Davey Smith G. Improving the accuracy of two-sample summary-data Mendelian

randomization: moving beyond the NOME assumption. Int J Epidemiol. 2019 Jun 1;48(3):728–742. doi: 10.1093/ije/dyy258.

Bowden J, Davey Smith G, Haycock PC, Burgess S. Consistent Estimation in Mendelian Randomization with Some Invalid Instruments Using a Weighted Median Estimator. Genet Epidemiol. 2016 May;40(4):304–14. doi: 10.1002/gepi.21965.

Bowden J, Davey Smith G, Burgess S. Mendelian randomization with invalid instruments: effect estimation and bias detection through Egger regression. Int J Epidemiol. 2015 Apr;44(2):512–25. doi: 10.1093/ije/dyv080.

Brown RH, Al-Chalabi A. Amyotrophic Lateral Sclerosis. N Engl J Med. 2017 Jul 13;377(2):162-172. doi: 10.1056/NEJMra1603471.

Cereda C, Baiocchi C, Bongioanni P, Cova E, Guareschi S, Metelli MR, Rossi B, Sbalsi I, Cuccia MC, Ceroni M. TNF and sTNFR1/2 plasma levels in ALS patients. J Neuroimmunol. 2008 Feb;194(1-2):123–31. doi: 10.1016/j.jneuroim.2007.10.028.

Chia R, Moaddel R, Kwan JY, Rasheed M, Ruffo P, Landeck N, Reho P, Vasta R, Calvo A, Moglia C, Canosa A, Manera U, Snyder A, Saez-Atienzar S, Grassano M, Brunetti M, Casale F, Ray A, Arvind K, Comertpay B, Zhu M, Gibbs JR; American Genome Center; Alba C, Dawson TM, Rosenthal LS, Hall AJ, Pantelyat AY, Narendra DP, Ehrlich DJ, Walker KA, Kosa P, Bielekova B, Egan JM, Candia J, Tanaka T, Ferrucci L, Dalgard CL, Scholz SW, Chiò A, Traynor BJ. A plasma proteomics-based candidate biomarker panel predictive of amyotrophic lateral sclerosis. Nat Med. 2025 Oct;31(10):3440–3450. doi: 10.1038/s41591-025-03890-6.

Chiò A, Calvo A, Moglia C, Mazzini L, Mora G; PARALS study group. Phenotypic heterogeneity of amyotrophic lateral sclerosis: a population based study. J Neurol Neurosurg Psychiatry. 2011 Jul;82(7):740–6. doi: 10.1136/jnnp.2010.235952.

Dellar ER, Vendrell I, Talbot K, Kessler BM, Fischer R, Turner MR, Thompson AG. Data-independent acquisition proteomics of cerebrospinal fluid implicates endoplasmic reticulum and inflammatory mechanisms in amyotrophic lateral sclerosis. J Neurochem. 2024 Feb;168(2):115–127. doi: 10.1111/jnc.16030.

De Houwer JFH, Dopper EG, van Buuren R, Stokkel M, de Boer L, Swartenbroekx T, Boesjes PA, Rajicic A, Sogorb-Esteve A, Bouzigues A, Russell LL, Foster PH, Ferry-Bolder E, van Swieten JC, Jiskoot LC, Sanchez-Valle R, Laforce R, Graff C, Galimberti D, Vandenberghe R, de Mendonça A, Tiraboschi P, Santana I, Gerhard A, Levin J, Nacmias B, Otto M, Bertoux M, Lebouvier T, Ducharme S, Butler CR, Le Ber I, Finger E, Tartaglia MC, Masellis M, Rowe JB, Synofzik M, Moreno F, Borroni B, Zetterberg H, Rohrer JD, Tijms BM, Pijnenburg YAL, Teunissen C, Seelaar H; GENFI Consortium Members. Distinct proteomic CSF profiles in genetic frontotemporal lobar degeneration. Brain. 2026 Aug 3;149(8):2816–2830. doi: 10.1093/brain/awaf457. PMID: 41343108; PMCID: PMC13431664.

Firth D, Bias reduction of maximum likelihood estimates, Biometrika. 1993;80(1):27–38. doi: 10.1093/biomet/80.1.27.

Fogh I, Lin K, Tiloca C, Rooney J, Gellera C, Diekstra FP, Ratti A, Shatunov A, van Es MA, Proitsi P, Jones A, Sproviero W, Chiò A, McLaughlin RL, Sorarù G, Corrado L, Stahl D, Del Bo R, Cereda C, Castellotti B, Glass JD, Newhouse S, Dobson R, Smith BN, Topp S, van Rheenen W, Meininger V, Melki J, Morrison KE, Shaw PJ, Leigh PN, Andersen PM, Comi GP, Ticozzi N, Mazzini L, D’Alfonso S, Traynor BJ, Van Damme P, Robberecht W, Brown RH, Landers JE, Hardiman O, Lewis CM, van den Berg LH, Shaw CE, Veldink JH, Silani V, Al-Chalabi A, Powell J. Association of a Locus in the CAMTA1 Gene With Survival in Patients With Sporadic Amyotrophic Lateral Sclerosis. JAMA Neurol. 2016 Jul 1;73(7):812–20. doi: 10.1001/jamaneurol.2016.1114.

Gaiani A, Martinelli I, Bello L, Querin G, Puthenparampil M, Ruggero S, Toffanin E, Cagnin A, Briani C, Pegoraro E, Sorarù G. Diagnostic and Prognostic Biomarkers in Amyotrophic Lateral Sclerosis: Neurofilament Light Chain Levels in Definite Subtypes of Disease. JAMA Neurol. 2017 May 1;74(5):525–532. doi: 10.1001/jamaneurol.2016.5398.

Gao J, Dharmadasa T, Malaspina A, Shaw PJ, Talbot K, Turner MR, Thompson AG. Creatine kinase and prognosis in amyotrophic lateral sclerosis: a literature review and multi-centre cohort analysis. J Neurol. 2022 Oct;269(10):5395–5404. doi: 10.1007/s00415-022-11195-8.

Genomics England. Adult onset neurodegenerative disorder v7.5. PanelApp. Available: https://panelapp.genomicsengland.co.uk/panels/474 [Accessed 08 Sep 2025].

Giambartolomei C, Vukcevic D, Schadt EE, Franke L, Hingorani AD, Wallace C, Plagnol V. Bayesian test for colocalisation between pairs of genetic association studies using summary statistics. PLoS Genet. 2014 May 15;10(5):e1004383. doi: 10.1371/journal.pgen.1004383.

Guidotti G, Scarlata C, Brambilla L, Rossi D. Tumor Necrosis Factor Alpha in Amyotrophic Lateral Sclerosis: Friend or Foe? Cells. 2021 Mar 1;10(3):518. doi: 10.3390/cells10030518.

Hensley K, Fedynyshyn J, Ferrell S, Floyd RA, Gordon B, Grammas P, Hamdheydari L, Mhatre M, Mou S, Pye QN, Stewart C, West M, West S, Williamson KS. Message and protein-level elevation of tumor necrosis factor alpha (TNF alpha) and TNF alpha-modulating cytokines in spinal cords of the G93A-SOD1 mouse model for amyotrophic lateral sclerosis. Neurobiol Dis. 2003 Oct;14(1):74–80. doi: 10.1016/s0969-9961(03)00087-1.

Iizuka N, Oka M, Yamada-Okabe H, Nishida M, Maeda Y, Mori N, Takao T, Tamesa T, Tangoku A, Tabuchi H, Hamada K, Nakayama H, Ishitsuka H, Miyamoto T, Hirabayashi A, Uchimura S, Hamamoto Y. Oligonucleotide microarray for prediction of early intrahepatic recurrence of hepatocellular carcinoma after curative resection. Lancet. 2003 Mar 15;361(9361):923–9. doi: 10.1016/S0140-6736(03)12775-4.

Imam F, Saloner R, Vogel JW, Krish V, Abdel-Azim G, Ali M, An L, Anastasi F, Bennett D, Pichet Binette A, Boxer AL, Bringmann M, Burns JM, Cruchaga C, Dage JL, Farinas A, Ferrucci L, Finney CA, Frasier M, Hansson O, Hohman TJ, Johnson ECB, Kivimaki M, Korologou-Linden R, Ruiz Laza A, Levey AI, Liepelt-Scarfone I, Lu L, Mattsson-Carlgren N, Middleton LT, Nho K, Oh HS, Petersen RC, Reiman EM, Robinson O, Rothstein JD, Saykin AJ, Shvetcov A, Slawson C, Smets B, Suárez-Calvet M, Tijms BM, Timmers M, Vieira F, Vilor-Tejedor N, Visser PJ, Walker KA, Winchester LM, Wyss-Coray T, Yang C, Bose N, Lovestone S; Global Neurodegeneration Proteomics Consortium (GNPC). The Global Neurodegeneration Proteomics Consortium: biomarker and drug target discovery for common neurodegenerative diseases and aging. Nat Med. 2025 Aug;31(8):2556–2566. doi: 10.1038/s41591-025-03834-0.

Jaganathan K, Kyriazopoulou Panagiotopoulou S, McRae JF, Darbandi SF, Knowles D, Li YI, Kosmicki JA, Arbelaez J, Cui W, Schwartz GB, Chow ED, Kanterakis E, Gao H, Kia A, Batzoglou S, Sanders SJ, Farh KK. Predicting Splicing from Primary Sequence with Deep Learning. Cell. 2019 Jan 24;176(3):535–548.e24. doi: 10.1016/j.cell.2018.12.015.

Kia A, McAvoy K, Krishnamurthy K, Trotti D, Pasinelli P. Astrocytes expressing ALS-linked mutant FUS induce motor neuron death through release of tumor necrosis factor-alpha. Glia. 2018 May;66(5):1016–1033. doi: 10.1002/glia.23298.

Kim WJ, Chung M, Asress S, Trautwig AN, Dammer E, Fournier CN, Seyfried NT, McEachin ZT, Glass JD, Thomas EV. Ten protein panel predicts ALS from plasma with high accuracy. Nat Commun. 2026 Oct 19;17(1):11768. doi: 10.1038/s41467-026-78766-5. PMID: 43045712; PMCID: PMC13778276.

Kircher M, Witten DM, Jain P, O’Roak BJ, Cooper GM, Shendure J. A general framework for estimating the relative pathogenicity of human genetic variants. Nat Genet. 2014 Mar;46(3):310–5. doi: 10.1038/ng.2892.

Klimovski H, Weinreich M, Strange A, Magen I, Yacovzada NS, Cohen Y, Melamed Kadosh D, Lester DG, Taylor A, Zhou Y, Ziv T, Abramovich B, Subramaniam AG, Perlson E, Drory V, Admon A, Shaw PJ, Malaspina A, Turner MR, Talbot K, Thompson AG, Cooper-Knock J, Hornstein E. Circulating protein profiling identifies prognostic biomarkers in amyotrophic lateral sclerosis. medRxiv. 2026 Jul 23:2026–07.

Lawlor DA, Harbord RM, Sterne JA, Timpson N, Davey Smith G. Mendelian randomization: using genes as instruments for making causal inferences in epidemiology. Stat Med. 2008 Apr 15;27(8):1133–63. doi: 10.1002/sim.3034.

Lee S, Emond MJ, Bamshad MJ, Barnes KC, Rieder MJ, Nickerson DA; NHLBI GO Exome Sequencing Project—ESP Lung Project Team; Christiani DC, Wurfel MM, Lin X. Optimal unified approach for rare-variant association testing with application to small-sample case-control whole-exome sequencing studies. Am J Hum Genet. 2012 Aug 10;91(2):224–37. doi: 10.1016/j.ajhg.2012.06.007.

Marttila M, Lehtokari VL, Marston S, Nyman TA, Barnerias C, Beggs AH, Bertini E, Ceyhan-Birsoy O, Cintas P, Gerard M, Gilbert-Dussardier B, Hogue JS, Longman C, Eymard B, Frydman M, Kang PB, Klinge L, Kolski H, Lochmüller H, Magy L, Manel V, Mayer M, Mercuri E, North KN, Peudenier-Robert S, Pihko H, Probst FJ, Reisin R, Stewart W, Taratuto AL, de Visser M, Wilichowski E, Winer J, Nowak K, Laing NG, Winder TL, Monnier N, Clarke NF, Pelin K, Grönholm M, Wallgren-Pettersson C. Mutation update and genotype-phenotype correlations of novel and previously described mutations in TPM2 and TPM3 causing congenital myopathies. Hum Mutat. 2014 Jul;35(7):779–90. doi: 10.1002/humu.22554.

Project MinE ALS Sequencing Consortium. Project MinE: study design and pilot analyses of a large-scale whole-genome sequencing study in amyotrophic lateral sclerosis. Eur J Hum Genet. 2018 Oct;26(10):1537–1546. doi: 10.1038/s41431-018-0177-4.

McLaren W, Pritchard B, Rios D, Chen Y, Flicek P, Cunningham F. Deriving the consequences of genomic variants with the Ensembl API and SNP Effect Predictor. Bioinformatics. 2010 Aug 15;26(16):2069–70. doi: 10.1093/bioinformatics/btq330.

Miller RG, Mitchell JD, Moore DH. Riluzole for amyotrophic lateral sclerosis (ALS)/motor neuron disease (MND). Cochrane Database Syst Rev. 2012 Mar 14;2012(3):CD001447. doi: 10.1002/14651858.CD001447.pub3.

Mitsumoto H, Brooks BR, Silani V. Clinical trials in amyotrophic lateral sclerosis: why so many negative trials and how can trials be improved? Lancet Neurol. 2014 Nov;13(11):1127–1138. doi: 10.1016/S1474-4422(14)70129-2.

Ran X, Wuu J, Qin ZS, Cooper-Knock J, Granit V, Grignon AL, Li Y, Lin E, Fernandez MC, Colato D, Carberry N, Lill CM, Piazza P, Malaspina A, Benatar M. Predicting Phenoconversion to Clinically Manifest ALS: Results of a Large-Scale Proteomic Study. MedRxiv. Published online 8^th^ December 2025. doi: 10.64898/2025.12.06.25341403.

Reches M, Porat Y, Gazit E. Amyloid fibril formation by pentapeptide and tetrapeptide fragments of human calcitonin. J Biol Chem. 2002 Sep 20;277(38):35475–80. doi: 10.1074/jbc.M206039200.

Rosenfeld MG, Mermod JJ, Amara SG, Swanson LW, Sawchenko PE, Rivier J, Vale WW, Evans RM. Production of a novel neuropeptide encoded by the calcitonin gene via tissue-specific RNA processing. Nature. 1983 Jul 14-20;304(5922):129–35. doi: 10.1038/304129a0.

Royston P, Altman DG. External validation of a Cox prognostic model: principles and methods. BMC Med Res Methodol. 2013 Mar 6;13:33. doi: 10.1186/1471-2288-13-33.

Rubin DB, Multiple Imputation for Nonresponse in Surveys (John Wiley and Sons, 1987).

Russo AF, Hay DL. CGRP physiology, pharmacology, and therapeutic targets: migraine and beyond. Physiol Rev. 2023 Apr 1;103(2):1565–1644. doi: 10.1152/physrev.00059.2021. Epub 2022 Dec 1. PMID: 36454715; PMCID: PMC9988538.

Sabbatini D, Raggi F, Ruggero S, Seguso M, Mandrioli J, Cagnin A, Briani C, Toffanin E, Gizzi M, Fortuna A, Bello L, Pegoraro E, Musso G, Sorarù G. Evaluation of peripherin in biofluids of patients with motor neuron diseases. Ann Clin Transl Neurol. 2021 Aug;8(8):1750–1754. doi: 10.1002/acn3.51419.

Sinha SK, Zachariah S, Quiñones HI, Shindo M, Chaudhary PM. Role of TRAF3 and -6 in the activation of the NF-kappa B and JNK pathways by X-linked ectodermal dysplasia receptor. J Biol Chem. 2002 Nov 22;277(47):44953–61. doi: 10.1074/jbc.M207923200.

Stommel EW, Cohen JA, Fadul CE, Cogbill CH, Graber DJ, Kingman L, Mackenzie T, Channon Smith JY, Harris BT. Efficacy of thalidomide for the treatment of amyotrophic lateral sclerosis: a phase II open label clinical trial. Amyotroph Lateral Scler. 2009 Oct-Dec;10(5-6):393–404. doi: 10.3109/17482960802709416.

Strange A, Weinreich M, Klimovski H, Holdom C, Daniel A, Baidoun M, Mahiddine F, Wong K, Magen I, Chen Y, Turner M, Talbot K, Lester DG, Thompson AG, Hornstein E, Zhang S, Cooper-Knock J, Shaw PJ. Circulating gelsolin tracks disease stage in amyotrophic lateral sclerosis across four cohorts. medRxiv 2026.11.18.26368465; doi: 10.64898/2026.11.18.26368465.

Sun BB, Chiou J, Traylor M, Benner C, Hsu YH, Richardson TG, Surendran P, Mahajan A, Robins C, Vasquez-Grinnell SG, Hou L, Kvikstad EM, Burren OS, Davitte J, Ferber KL, Gillies CE, Hedman ÅK, Hu S, Lin T, Mikkilineni R, Pendergrass RK, Pickering C, Prins B, Baird D, Chen CY, Ward LD, Deaton AM, Welsh S, Willis CM, Lehner N, Arnold M, Wörheide MA, Suhre K, Kastenmüller G, Sethi A, Cule M, Raj A; Alnylam Human Genetics; AstraZeneca Genomics Initiative; Biogen Biobank Team; Bristol Myers Squibb; Genentech Human Genetics; GlaxoSmithKline Genomic Sciences; Pfizer Integrative Biology; Population Analytics of Janssen Data Sciences; Regeneron Genetics Center; Burkitt-Gray L, Melamud E, Black MH, Fauman EB, Howson JMM, Kang HM, McCarthy MI, Nioi P, Petrovski S, Scott RA, Smith EN, Szalma S, Waterworth DM, Mitnaul LJ, Szustakowski JD, Gibson BW, Miller MR, Whelan CD. Plasma proteomic associations with genetics and health in the UK Biobank. Nature. 2023 Oct;622(7982):329–338. doi: 10.1038/s41586-023-06592-6.

Taylor A, Fletcher MP. easyplater: The easy way to generate microplate designs deconvolved from multivariate clinical data. arXiv. 2025 Dec; doi: 10.48550/arXiv.2512.17988.

Teunissen CE, Petzold A, Bennett JL, Berven FS, Brundin L, Comabella M, Franciotta D, Frederiksen JL, Fleming JO, Furlan R, Hintzen RQ, Hughes SG, Johnson MH, Krasulova E, Kuhle J, Magnone MC, Rajda C, Rejdak K, Schmidt HK, van Pesch V, Waubant E, Wolf C, Giovannoni G, Hemmer B, Tumani H, Deisenhammer F. A consensus protocol for the standardization of cerebrospinal fluid collection and biobanking. Neurology. 2009 Dec 1;73(22):1914–22. doi: 10.1212/WNL.0b013e3181c47cc2.

Thompson AG, Gray E, Verber N, Bobeva Y, Lombardi V, Shepheard SR, Yildiz O, Feneberg E, Farrimond L, Dharmadasa T, Gray P, Edmond EC, Scaber J, Gagliardi D, Kirby J, Jenkins TM, Fratta P, McDermott CJ, Manohar SG, Talbot K, Malaspina A, Shaw PJ, Turner MR. Multicentre appraisal of amyotrophic lateral sclerosis biofluid biomarkers shows primacy of blood neurofilament light chain. Brain Commun. 2022 Feb 9;4(1):fcac029. doi: 10.1093/braincomms/fcac029.

Tibshirani R. Regression Shrinkage and Selection via the Lasso. J R Stat Soc Ser B Methodol. 1996;58(1):267–88.

Trautwig AN, Fox EJ, Dammer EB, Shantaraman A, Ping L, Duong DM, Watson CM, Wu F, Asress S, Guo Q, Levey AI, Lah JJ, Verde F, Doretti A, Ratti A, Ticozzi N, Ly CV, Miller TM, Garret MA, Berry JD, Thomas EV, Fournier CN, McEachin ZT, Seyfried NT, Glass JD. Network analysis of the cerebrospinal fluid proteome reveals shared and unique differences between sporadic and familial forms of amyotrophic lateral sclerosis. Mol Neurodegener. 2025 May 15;20(1):58. doi: 10.1186/s13024-025-00838-9.

Van Den Berg LH, Sorenson E, Gronseth G, Macklin EA, Andrews J, Baloh RH, Benatar M, Berry JD, Chio A, Corcia P, Genge A, Gubitz AK, Lomen-Hoerth C, McDermott CJ, Pioro EP, Rosenfeld J, Silani V, Turner MR, Weber M, Brooks BR, Miller RG, Mitsumoto H; Airlie House ALS Clinical Trials Guidelines Group. Revised Airlie House consensus guidelines for design and implementation of ALS clinical trials. Neurology. 2019 Apr 2;92(14):e1610–e1623. doi: 10.1212/WNL.0000000000007242.

van Eijk RPA, de Jongh AD, Nikolakopoulos S, McDermott CJ, Eijkemans MJC, Roes KCB, van den Berg LH. An old friend who has overstayed their welcome: the ALSFRS-R total score as primary endpoint for ALS clinical trials. Amyotroph Lateral Scler Frontotemporal Degener. 2021 May;22(3-4):300–307. doi: 10.1080/21678421.2021.1879865.

van Rheenen W, van der Spek RAA, Bakker MK, van Vugt JJFA, Hop PJ, Zwamborn RAJ, de Klein N, Westra HJ, Bakker OB, Deelen P, Shireby G, Hannon E, Moisse M, Baird D, Restuadi R, Dolzhenko E, Dekker AM, Gawor K, Westeneng HJ, Tazelaar GHP, van Eijk KR, Kooyman M, Byrne RP, Doherty M, Heverin M, Al Khleifat A, Iacoangeli A, Shatunov A, Ticozzi N, Cooper-Knock J, Smith BN, Gromicho M, Chandran S, Pal S, Morrison KE, Shaw PJ, Hardy J, Orrell RW, Sendtner M, Meyer T, Başak N, van der Kooi AJ, Ratti A, Fogh I, Gellera C, Lauria G, Corti S, Cereda C, Sproviero D, D’Alfonso S, Sorarù G, Siciliano G, Filosto M, Padovani A, Chiò A, Calvo A, Moglia C, Brunetti M, Canosa A, Grassano M, Beghi E, Pupillo E, Logroscino G, Nefussy B, Osmanovic A, Nordin A, Lerner Y, Zabari M, Gotkine M, Baloh RH, Bell S, Vourc’h P, Corcia P, Couratier P, Millecamps S, Meininger V, Salachas F, Mora Pardina JS, Assialioui A, Rojas-García R, Dion PA, Ross JP, Ludolph AC, Weishaupt JH, Brenner D, Freischmidt A, Bensimon G, Brice A, Durr A, Payan CAM, Saker-Delye S, Wood NW, Topp S, Rademakers R, Tittmann L, Lieb W, Franke A, Ripke S, Braun A, Kraft J, Whiteman DC, Olsen CM, Uitterlinden AG, Hofman A, Rietschel M, Cichon S, Nöthen MM, Amouyel P; SLALOM Consortium; PARALS Consortium; SLAGEN Consortium; SLAP Consortium; Traynor BJ, Singleton AB, Mitne Neto M, Cauchi RJ, Ophoff RA, Wiedau-Pazos M, Lomen-Hoerth C, van Deerlin VM, Grosskreutz J, Roediger A, Gaur N, Jörk A, Barthel T, Theele E, Ilse B, Stubendorff B, Witte OW, Steinbach R, Hübner CA, Graff C, Brylev L, Fominykh V, Demeshonok V, Ataulina A, Rogelj B, Koritnik B, Zidar J, Ravnik-Glavač M, Glavač D, Stević Z, Drory V, Povedano M, Blair IP, Kiernan MC, Benyamin B, Henderson RD, Furlong S, Mathers S, McCombe PA, Needham M, Ngo ST, Nicholson GA, Pamphlett R, Rowe DB, Steyn FJ, Williams KL, Mather KA, Sachdev PS, Henders AK, Wallace L, de Carvalho M, Pinto S, Petri S, Weber M, Rouleau GA, Silani V, Curtis CJ, Breen G, Glass JD, Brown RH Jr, Landers JE, Shaw CE, Andersen PM, Groen EJN, van Es MA, Pasterkamp RJ, Fan D, Garton FC, McRae AF, Davey Smith G, Gaunt TR, Eberle MA, Mill J, McLaughlin RL, Hardiman O, Kenna KP, Wray NR, Tsai E, Runz H, Franke L, Al-Chalabi A, Van Damme P, van den Berg LH, Veldink JH. Common and rare variant association analyses in amyotrophic lateral sclerosis identify 15 risk loci with distinct genetic architectures and neuron-specific biology. Nat Genet. 2021 Dec;53(12):1636–1648. doi: 10.1038/s41588-021-00973-1.

Van Rheenen W, Shatunov A, Dekker AM, McLaughlin RL, Diekstra FP, Pulit SL, van der Spek RA, Võsa U, de Jong S, Robinson MR, Yang J, Fogh I, van Doormaal PT, Tazelaar GH, Koppers M, Blokhuis AM, Sproviero W, Jones AR, Kenna KP, van Eijk KR, Harschnitz O, Schellevis RD, Brands WJ, Medic J, Menelaou A, Vajda A, Ticozzi N, Lin K, Rogelj B, Vrabec K, Ravnik-Glavač M, Koritnik B, Zidar J, Leonardis L, Grošelj LD, Millecamps S, Salachas F, Meininger V, de Carvalho M, Pinto S, Mora JS, Rojas-García R, Polak M, Chandran S, Colville S, Swingler R, Morrison KE, Shaw PJ, Hardy J, Orrell RW, Pittman A, Sidle K, Fratta P, Malaspina A, Topp S, Petri S, Abdulla S, Drepper C, Sendtner M, Meyer T, Ophoff RA, Staats KA, Wiedau-Pazos M, Lomen-Hoerth C, Van Deerlin VM, Trojanowski JQ, Elman L, McCluskey L, Basak AN, Tunca C, Hamzeiy H, Parman Y, Meitinger T, Lichtner P, Radivojkov-Blagojevic M, Andres CR, Maurel C, Bensimon G, Landwehrmeyer B, Brice A, Payan CA, Saker-Delye S, Dürr A, Wood NW, Tittmann L, Lieb W, Franke A, Rietschel M, Cichon S, Nöthen MM, Amouyel P, Tzourio C, Dartigues JF, Uitterlinden AG, Rivadeneira F, Estrada K, Hofman A, Curtis C, Blauw HM, van der Kooi AJ, de Visser M, Goris A, Weber M, Shaw CE, Smith BN, Pansarasa O, Cereda C, Del Bo R, Comi GP, D’Alfonso S, Bertolin C, Sorarù G, Mazzini L, Pensato V, Gellera C, Tiloca C, Ratti A, Calvo A, Moglia C, Brunetti M, Arcuti S, Capozzo R, Zecca C, Lunetta C, Penco S, Riva N, Padovani A, Filosto M, Muller B, Stuit RJ; PARALS Registry; SLALOM Group; SLAP Registry; FALS Sequencing Consortium; SLAGEN Consortium; NNIPPS Study Group; Blair I, Zhang K, McCann EP, Fifita JA, Nicholson GA, Rowe DB, Pamphlett R, Kiernan MC, Grosskreutz J, Witte OW, Ringer T, Prell T, Stubendorff B, Kurth I, Hübner CA, Leigh PN, Casale F, Chio A, Beghi E, Pupillo E, Tortelli R, Logroscino G, Powell J, Ludolph AC, Weishaupt JH, Robberecht W, Van Damme P, Franke L, Pers TH, Brown RH, Glass JD, Landers JE, Hardiman O, Andersen PM, Corcia P, Vourc’h P, Silani V, Wray NR, Visscher PM, de Bakker PI, van Es MA, Pasterkamp RJ, Lewis CM, Breen G, Al-Chalabi A, van den Berg LH, Veldink JH. Genome-wide association analyses identify new risk variants and the genetic architecture of amyotrophic lateral sclerosis. Nat Genet. 2016 Sep;48(9):1043–8. doi: 10.1038/ng.3622.

Varma S, Simon R. Bias in error estimation when using cross-validation for model selection. BMC Bioinformatics. 2006 Feb 23;7:91. doi: 10.1186/1471-2105-7-91.

Westeneng HJ, Debray TPA, Visser AE, van Eijk RPA, Rooney JPK, Calvo A, Martin S, McDermott CJ, Thompson AG, Pinto S, Kobeleva X, Rosenbohm A, Stubendorff B, Sommer H, Middelkoop BM, Dekker AM, van Vugt JJFA, van Rheenen W, Vajda A, Heverin M, Kazoka M, Hollinger H, Gromicho M, Körner S, Ringer TM, Rödiger A, Gunkel A, Shaw CE, Bredenoord AL, van Es MA, Corcia P, Couratier P, Weber M, Grosskreutz J, Ludolph AC, Petri S, de Carvalho M, Van Damme P, Talbot K, Turner MR, Shaw PJ, Al-Chalabi A, Chiò A, Hardiman O, Moons KGM, Veldink JH, van den Berg LH. Prognosis for patients with amyotrophic lateral sclerosis: development and validation of a personalised prediction model. Lancet Neurol. 2018 May;17(5):423–433. doi: 10.1016/S1474-4422(18)30089-9.

Wik L, Nordberg N, Broberg J, Björkesten J, Assarsson E, Henriksson S, Grundberg I, Pettersson E, Westerberg C, Liljeroth E, Falck A, Lundberg M. Proximity Extension Assay in Combination with Next-Generation Sequencing for High-throughput Proteome-wide Analysis. Mol Cell Proteomics. 2021;20:100168. doi: 10.1016/j.mcpro.2021.100168.

Zuber V, Grinberg NF, Gill D, Manipur I, Slob EAW, Patel A, Wallace C, Burgess S. Combining evidence from Mendelian randomization and colocalization: Review and comparison of approaches. Am J Hum Genet. 2022 May 5;109(5):767–782. doi: 10.1016/j.ajhg.2022.04.001.

