## Supplementary figures for "A proteomic signature for prognostic stratification in amyotrophic lateral sclerosis"

1.	CSF and serum PEA CV and LOD plots

2.	CSF and serum PEA PCA and NPX plots

3.	CSF and serum assay warning raw count density plots

4.	CSF and serum PEA PCA 1-10 summaries

5.	CSF PEA PCA biological and technical factor plots

6.	Serum PEA PCA biological and technical factor plots

7.	CSF PEA plate layouts by sample collection cohort

8.	Serum PEA plate layouts by sample collection cohort

9.	CSF and serum PEA plate Kaplan-Meier curves

10.	CSF sample collection cohort Kaplan-Meier curves

11.	Serum sample collection cohort Kaplan-Meier curves

12.	CSF and serum PEA-MSD NEFL comparisons

13.	Overlapping serum PEA and DIA-MS proteins

14.	External validation cohort Kaplan-Meier curve

15.	Serum lasso-selected proteins raw count density plots

16.	CSF-serum PEA comparisons for lasso-selected proteins

17.	Serum CALCA joint model diagnostic plots

18.	Serum EDA2R joint model diagnostic plots

19.	Serum NEFL joint model diagnostic plots

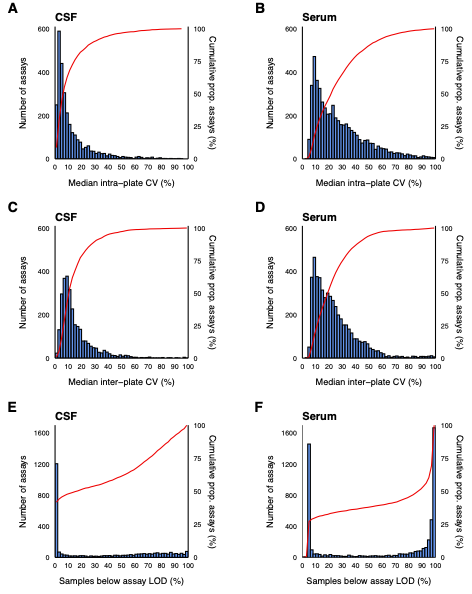

**Supplementary Figure 1**: Histograms representing median intra-plate assay coefficient of variation (CV) in (**A)** CSF and (**B)** serum, median inter-plate assay CVs in (**C**) CSF and (**D**) serum, and proportion of sample values below each assay’s limit of detection (LOD) in (**E**) CSF and (**F**) serum.

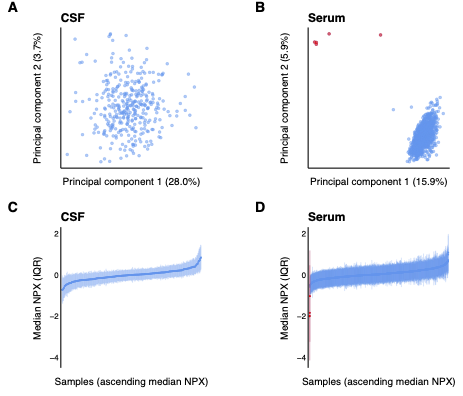

**Supplementary Figure 2**: Samples represented by the first two principal components in (**A**) CSF and (**B**) serum, and quartile boxplots representing sample Normalized Protein eXpression (NPX) in (**C**) CSF and (**D**) serum. The six serum samples removed during quality control are shown in red.

**CSF**

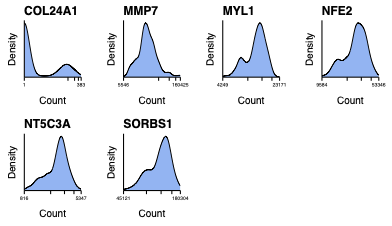

**Serum**

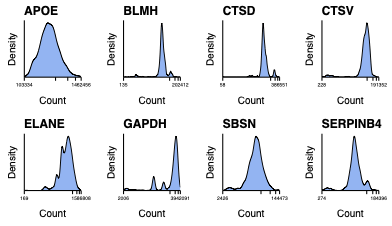

**Supplementary Figure 3**: Density plots of six CSF and eight serum assays with QC warnings. Minimum and maximum raw counts per assay across samples are given, together with interquartile and median count markers, on a log_10_ scale. All were retained for subsequent analyses.

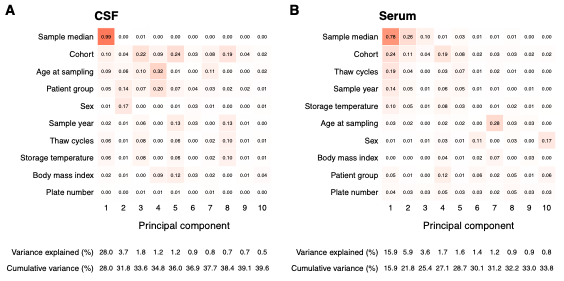

**Supplementary Figure 4**: Correlation plots of the coefficients of determination (*R*^2^) from linear regressions of selected technical and biological factors against PCs 1 to 10 in (**A**) CSF and (**B**) serum. Rows are hierarchically clustered in each plot. Shown with individual and cumulative proportion of explained variance.
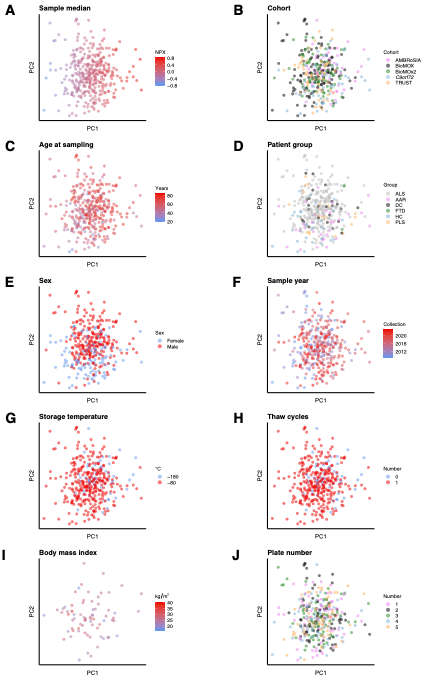

**Supplementary Figure 5**: Plots of technical and biological factors in *n*=356 CSF samples represented by their PC1 and PC2 values. Storage temperature and thaw cycles are fully collinear. Abbreviations: AAR – asymptomatic at risk; AMBRoSIA – A Multicentre Biomarker Resource Strategy in ALS; BioMOx – Oxford Study for Biomarkers in MND; *C9orf72* – C9orf72 Cohort Study; DC – disease control; FTD – frontotemporal dementia; HC – healthy control; PLS – primary lateral sclerosis; TRUST –Terazosin Repurposing Study in ALS.

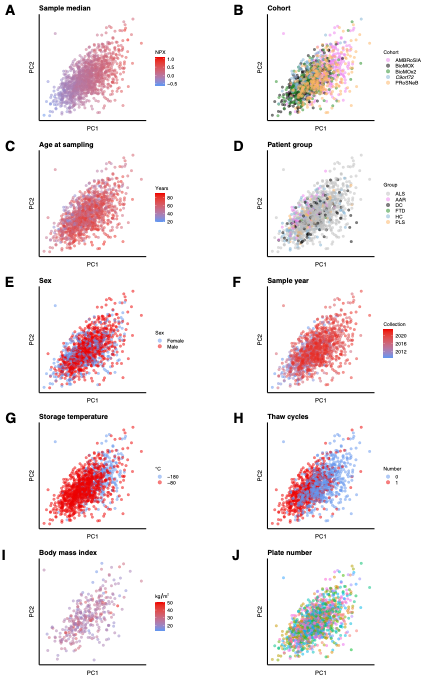

**Supplementary Figure 6**: Plots of technical and biological factors in *n*=1223 serum samples represented by their PC1 and PC2 values. 37 plate numbers not individually labelled. Abbreviations: ProSNeB – Prospective study of neurological biomarkers.

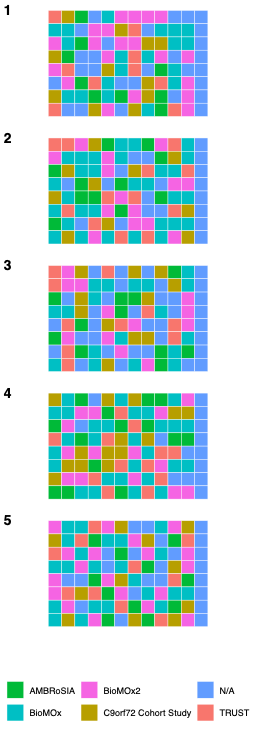

**Supplementary Figure 7**: CSF PEA 96-well plate maps by sample cohort.

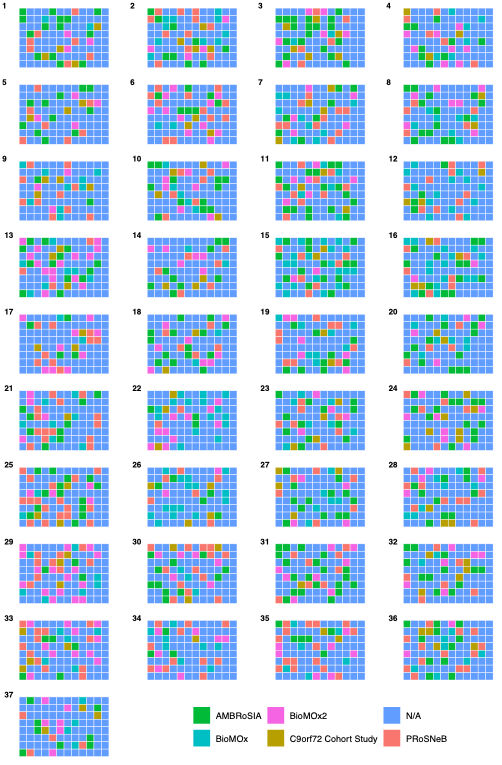

**Supplementary Figure 8**: Serum PEA 96-well plate maps by sample cohort.

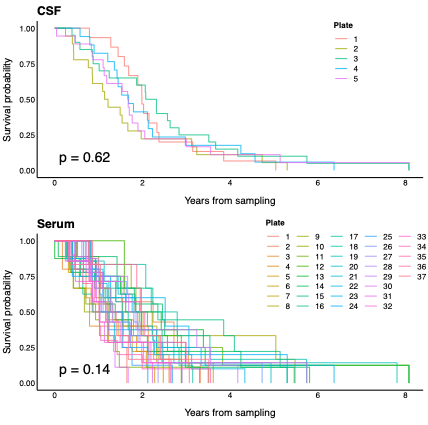

**Supplementary Figure 9**: Kaplan-Meier curves of time to death or censoring after sampling shown grouped by CSF and serum PEA plate number with log-rank *p*-value.

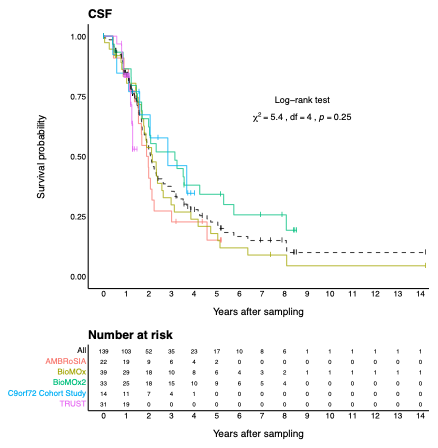

**Supplementary Figure 10**: Kaplan-Meier curves of survival after first-visit CSF sampling in patients with ALS for each sample collection cohorts and overall. Shown with risk table and omnibus log-rank result comparing survival between the cohorts.

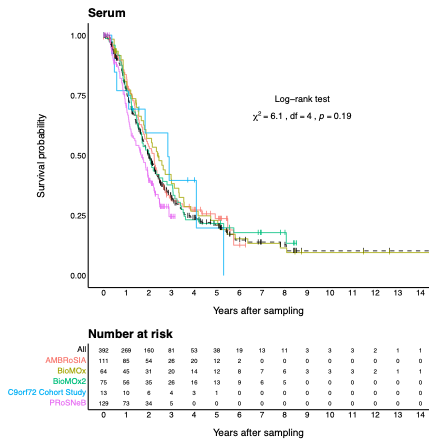

**Supplementary Figure 11**: Kaplan-Meier curves of survival after first-visit serum sampling in patients with ALS for each sample collection cohorts and overall. Shown with risk table and omnibus log-rank result comparing survival between the cohorts.

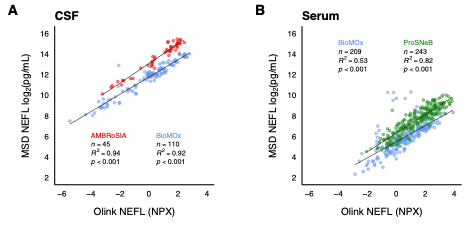

**Supplementary Figure 12**: Scatterplots showing the correlations between pre-existing MesoScale Discovery (MSD) electrochemiluminescence NEFL assay values and Olink PEA NEFL values from the same samples, measured in (**A**) CSF and (**B**) serum.
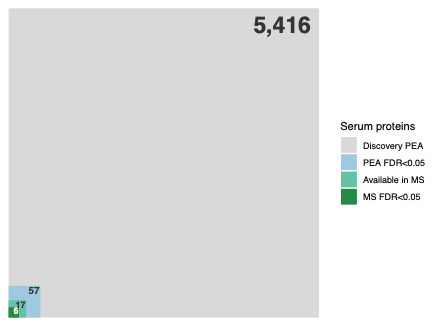

**Supplementary Figure 13**: Overview of the nested serum replication analysis to detect proteins with false discovery rate (FDR)-significant adjusted univariable Cox regression associations. Shown to scale.

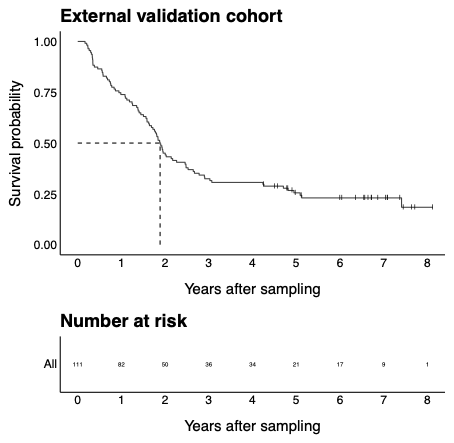
**Supplementary Figure 14**: Kaplan-Meier curve of time to death or censoring following serum sampling for patients in the Sheffield external validation cohort, shown with risk table. Median survival from sampling = 1.9 years (95% CI 1.7-2.5).

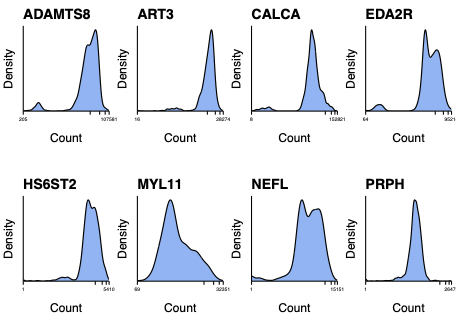

**Supplementary Figure 15**: Density plots of raw count values for the eight serum lasso-selected proteins, shown with minimum and maximum counts, and interquartile and median count markers on a log_10_ scale.

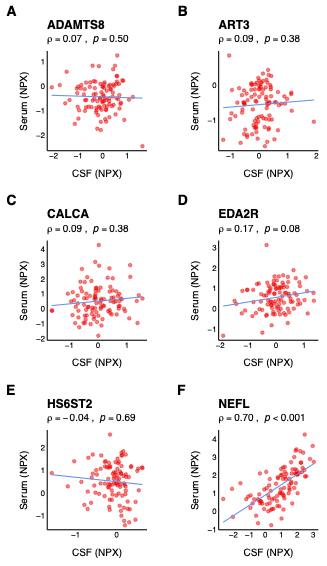

**Supplementary Figure 16**: Spearman’s rank correlation coefficients (⍴) of CSF and serum Olink NPX values for (**A**) a disintegrin and metallopeptidase with thrombospondin type 1 motif 8 (ADAMTS8), (**B**) ecto-ADP-ribosyltransferase 3 (ART3), (**C**) calcitonin (CALCA), (**D**) tumor necrosis factor receptor superfamily member 27 (EDA2R), (**E**) heparan sulfate 6-O-sulfotransferase 2 (HS6ST2) and (**F**) NEFL in *n*=104 patients with ALS who had both biofluids sampled at the same time.

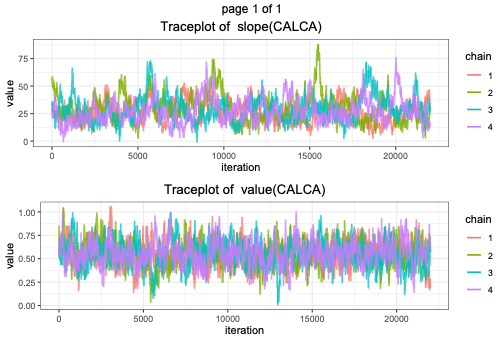

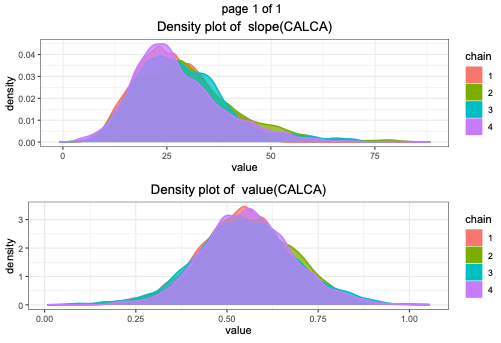

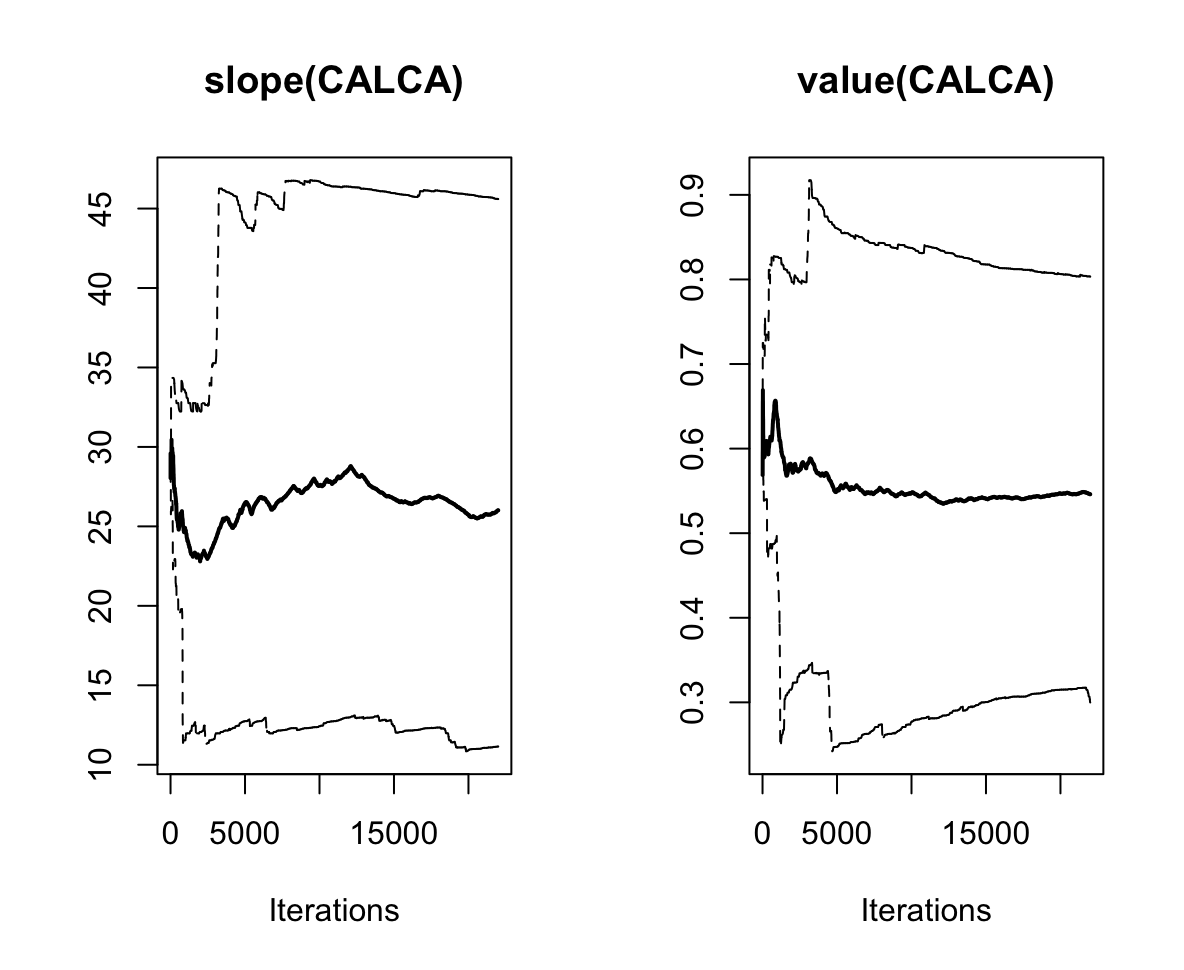

**Supplementary Figure 17**: Trace, density and cumulative quantile plots for the serum calcitonin (CALCA) joint model slope and value functional forms.

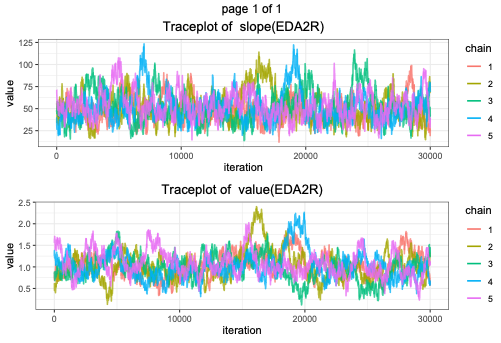

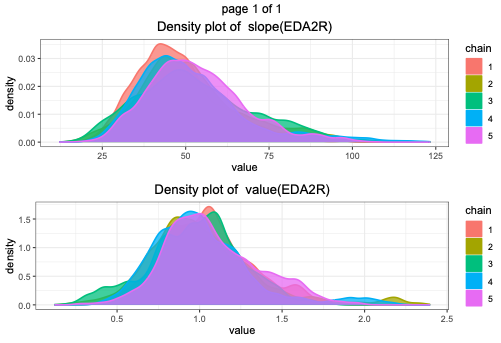

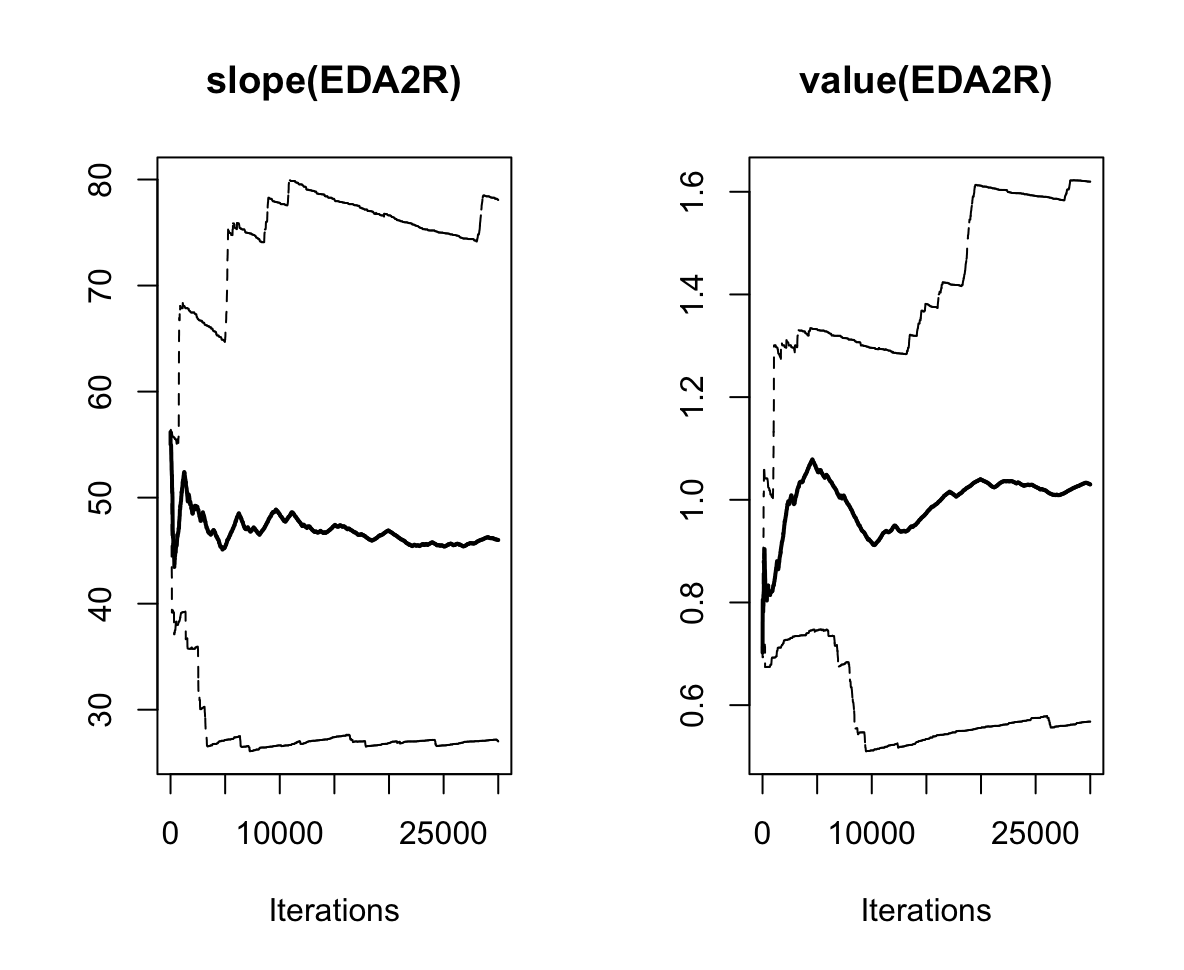

**Supplementary Figure 18:** Trace, density and cumulative quantile plots for the serum tumor necrosis factor receptor superfamily member 27 (EDA2R) joint model slope and value functional forms.

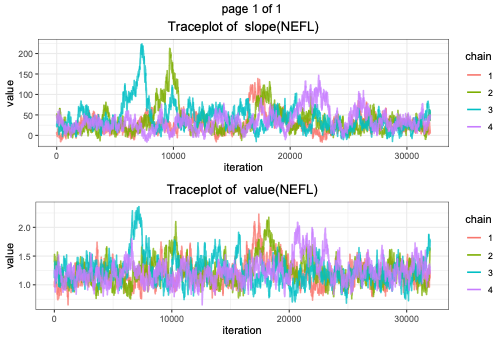

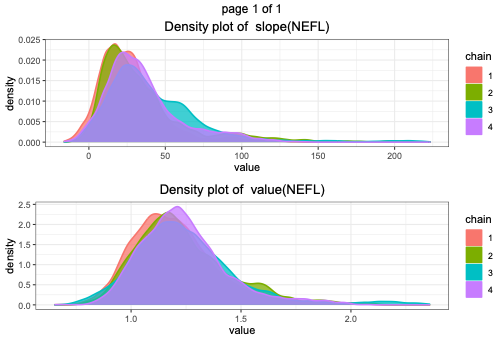

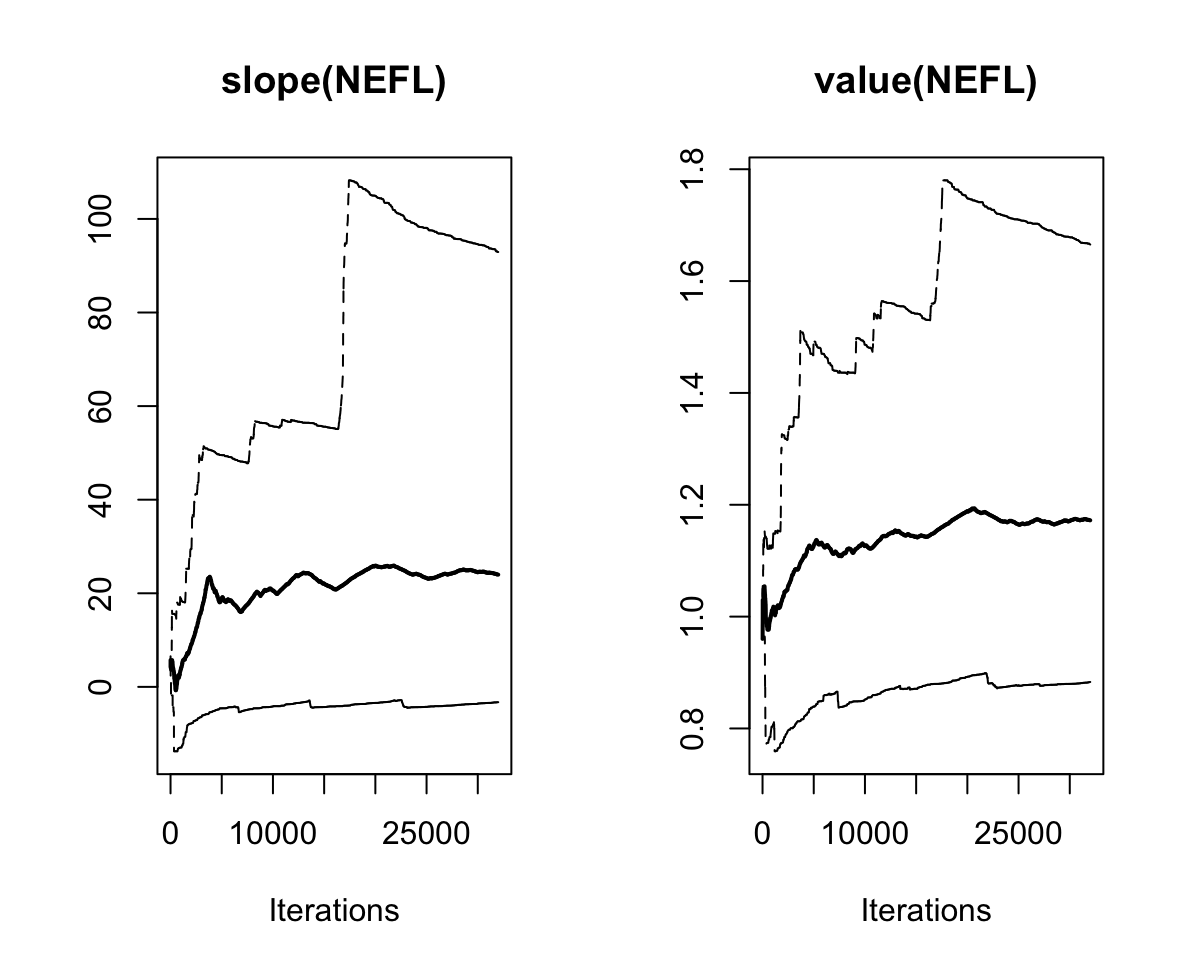

**Supplementary Figure 19:** Trace, density and cumulative quantile plots for the serum neurofilament light chain (NEFL) joint model slope and value functional forms.
